# Metagenomic analysis of the abundance and composition of antibiotic resistance genes in hospital wastewater in Benin, Burkina Faso, and Finland

**DOI:** 10.1101/2021.10.19.21265183

**Authors:** Melina A. Markkanen, Kaisa Haukka, Katariina M. M. Pärnänen, Victorien Tamegnon Dougnon, Isidore Juste O. Bonkoungou, Zakaria Garba, Halidou Tinto, Anniina Sarekoski, Antti Karkman, Anu Kantele, Marko P. J. Virta

**Affiliations:** Department of Microbiology, University of Helsinki, Finland; Multidisciplinary Center of Excellence in Antimicrobial Resistance Research, Finland; Research Unit in Applied Microbiology and Pharmacology of Natural Substances, Polytechnic School of Abomey-Calavi, University of Abomey-Calavi, Benin; Department of Biochemistry and Microbiology, University Joseph KI-ZERBO, Ouagadougou, Burkina Faso; Clinical Research Unit Nanoro, Institute for Research in Health Sciences, National Center for Scientific and Technological Research, Burkina Faso; Meilahti Infectious Diseases and Vaccine Research Center, MeiVac, Department of Infectious Diseases, University of Helsinki and Helsinki University Hospital, Helsinki, Finland; Human Microbiome Research Program, Faculty of Medicine, University of Helsinki, Helsinki, Finland

**Keywords:** antibiotic resistance, West Africa, hospital wastewater (HWW), carbapenemase, colistin resistance, metagenomes

## Abstract

Antibiotic resistance is one of the greatest global threats to human health, but substantial gaps in antibiotic resistance data exist in West African countries. We explored the presence of antibiotic resistance genes (ARGs) in the hospital wastewater (HWW) of nine hospitals in Benin and Burkina Faso and, for comparison, four hospitals in Finland using shotgun metagenomic sequencing. The highest sum of the relative abundance of ARGs in the 68 HWW samples was detected in Benin and the lowest in Finland. HWW resistomes and mobilomes in Benin and Burkina Faso resembled more of each other than those in Finland. Different carbapenemases were detected in varying abundances, especially in HWW from Burkina Faso and Finland. The *bla*GES genes, the most widespread carbapenemase genes in the Beninese HWW, were also found in water intended for hand-washing and in a puddle at the hospital yard in Benin. *mcr* were detected in the HWW of all three countries, with *mcr-5* being the most common *mcr* gene. These and other *mcr* genes were observed in very high relative abundances, even in treated wastewater in Burkina Faso and in a street gutter in Benin. The results provide evidence for public health decision-makers for the dire need to increase wastewater treatment capacity, with particular attention to HWW.

**Importance:** The global emergence and increased spread of antibiotic resistance threaten the effectiveness of antibiotics and, thus, the health of the entire population. Therefore, understanding the resistomes in different geographical locations is crucial in the global fight against the antibiotic crisis. However, this information is scarce in many low- and middle-income countries (LMICs), such as those in West Africa. In this study, we describe the resistomes of hospital wastewater in Benin and Burkina Faso and, as a comparison, in Finland. Our results help to understand the hitherto unrevealed resistance in Beninese and Burkinabe hospitals. Furthermore, the results emphasize the importance of wastewater management infrastructure design to minimize exposure events between humans, HWW, and the environment, preventing the circulation of resistant bacteria and ARGs between humans (hospitals and community) and the environment.

## 1. Introduction

The antibiotic resistance crisis is a global issue with multifaceted effects on human, animal, and environmental health, and it comes with substantial economic losses ^1^. Resistance to almost all antibiotics, including the last resort regimen reserved to treat the most severe infections, has emerged only a couple of years after introducing a new antibiotic to the global market ^2,3^. Due to limitations in diagnostic testing in low-resource settings in low- and middle-income countries (LMICs), broad-spectrum antibiotics are often prescribed empirically without microbiological verification of the causative pathogen or its sensitivity to different antibiotics ^4^. In addition, unregulated access to antibiotics allows their wide use for self-medication for humans and animals in these countries ^5–7^.

Acquired, potentially mobile ARGs are considered to have a pronounced clinical relevance and impact on the current AMR problem promoted by human activity ^8,9^. Furthermore, the genetic context of the ARG (e.g., whether under a strong promoter or not) influences its expression and the resulting resistance phenotype ^10^. Class 1 integrons are strongly linked to the dissemination of clinically relevant acquired ARGs ^11,12^. Although integrons are not mobile as such, multi-drug resistance gene cassettes carried in integrons can be transferred to new hosts, for example, via plasmids ^13^. The *intI1* and *qacEΔ* genes encoding the integron integrase and quaternary ammonium compound resistance, respectively, are typically used as markers for class 1 integrons ^12,14^.

As a consequence of the increased prevalence of extended-spectrum beta-lactamase-producing *Enterobacteriaceae* (ESBL-PE), the use of broad-spectrum antibiotics has increased in clinical practice ^2,15,16^, and attention has been directed to plasmid-mediated genes that encode carbapenemases. The carbapenemases *bla*GES, *bla*IMP, *bla*KPC, *bla*NDM, *bla*OXA-48, *bla*OXA-58, and *bla*VIM carried by plasmids have emerged and spread around the world during the past three decades ^17–20^. Colistin is a last-resort antibiotic used for treating infections caused by multi-drug and extensively resistant bacteria ^21^. The rapid emergence of colistin resistance mediated by *mcr* genes threatens the efficacy of colistin in clinical use ^22,23^.

Although AMR is of global concern, the crisis affects LMICs most dramatically, such as those in West Africa ^3,4,24–26^. Lack of research data is a major factor hindering the development of solutions to tackle the AMR problem in these countries ^4,24,27,28^. Despite gaps in resistance surveillance data, it is well known that the level of AMR is highest in many LMICs, such as in many African countries ^15,25,29,30^. By contrast, in Northern European countries, such as Finland, AMR occurrence is among the lowest globally, both in the community ^30,31^ and in healthcare settings ^32^.

As hospital wastewater (HWW) from healthcare facilities is considered to be at the frontline of AMR emergence and spread due to the heavy use of antibiotics and presence of immunocompromised patients ^33^, we set out to investigate the AMR situation and the characteristics of the resistomes in nine hospitals in two West African countries, Benin and Burkina Faso, where prior data was scarce ^15^. For comparison, we analyzed samples from four hospitals in Finland, where the level of AMR was expected to be low ^32^. As the HWW resistomes in Benin and Burkina Faso were largely unexplored targeted PCR-based methods ^34^, were not suitable for our study setting. Instead, in this study, we applied shotgun metagenomic sequencing to obtain a holistic view of the resistomes and mobilomes (a set of ARGs and MGEs, respectively) and the microbial communities present in the studied environment.

## 2. Results

### 2.1. General features of resistomes, mobilomes, and taxonomical compositions in HWW and other water samples from Benin, Burkina Faso, and Finland

We studied hospital wastewater (HWW) collected from Benin, Burkina Faso, and Finland (Figure S1, Table 1, Table S1-S2). On average, 31 million sequence reads per sample were analyzed. HWW from Benin showed the highest, and from Burkina Faso, the second-highest abundance of all detected ARGs normalized to bacterial 16S rRNA genes (sum of the relative abundance of ARGs). The sum of the relative abundance was the lowest in HWW from Finland (Figure 1A). Also the lowest diversity of ARGs was observed in HWW from Finland and the highest in Burkina Faso (Figure S3A).

**Table 1.** Sample information. HWW samples from hospitals in Benin (n = 26), Burkina Faso (n = 34), and Finland (n = 8) are denoted with a grey background, while the other samples are on white background.

| Sample ID | Country | Category | no. Samples in the Category | Hospital/Area | Collection Date | HWW |
| --- | --- | --- | --- | --- | --- | --- |
| BH01...BH09 | Benin | BENN HWW A | 7 | A | 27_11_19 | YES |
| BH27...BH39 | Benin | BENN HWW B | 11 | B | 29_11_19 | YES |
| BH44...BH50 | Benin | BENN HWW C | 5 | C | 09_12_19 | YES |
| BH58...BH61 | Benin | BENN HWW D | 3 | D | 11_12_19 | YES |
| BFH1...BFH4 | Burkina Faso | BF HWW E | 2 | E | 22_11_19 | YES |
| BFH6...BFH12 | Burkina Faso | BF HWW F | 6 | F | 28_11_19 | YES |
| BFH13...BFH15 | Burkina Faso | BF HWW G | 3 | G | 28_11_19 | YES |
| BFH16...BFH28 | Burkina Faso | BF HWW H | 10 | H | 04_12_19 | YES |
| BFH29...BFH41 | Burkina Faso | BF HWW I | 13 | I | 12_12_19 | YES |
| FH1 | Finland | FI HWW J | 1 | J | 20_01_20 | YES |
| FH2 | Finland | FI HWW K | 1 | K | 20_01_20 | YES |
| FH3 | Finland | FI HWW L | 1 | L | 20_01_20 | YES |
| FH4...FH6 | Finland | FI HWW M | 3 | M | 20_01_20 | YES |
| FH7 | Finland | FI HWW N | 1 | N | 23_01_20 | YES |
| FH9 | Finland | FI HWW O | 1 | O | 28_01_20 | YES |
| BH11 | Benin | BENN well water A (drinking) | 1 | A | 27_11_19 | NO |
| BH13 | Benin | BENN street gutter B | 1 | B | 27_11_19 | NO |
| BH48 | Benin | BENN puddle at yard C | 1 | C | 09_12_19 | NO |
| BH52 | Benin | BENN hand-washing C | 1 | C | 09_12_19 | NO |
| BSE100 | Benin | BENN river P (drinking) | 1 | P | 04_12_19 | NO |
| BSE74 | Benin | BENN river Q (drinking) | 1 | Q | 04_12_19 | NO |
| BSE79 | Benin | BENN river R (drinking) | 1 | R | 04_12_19 | NO |
| BSE93 | Benin | BENN tap water S (drinking) | 1 | S | 04_12_19 | NO |
| BFH26 | Burkina Faso | BF exit after biological treatment | 1 | H | 04_12_19 | NO |
| BFH27 | Burkina Faso | BF wetland receiving treated HWW | 1 | H | 04_12_19 | NO |
| BFH42 | Burkina Faso | BF receiving river after WWTP | 1 | T | 12_12_19 | NO |

**Figure 1.**
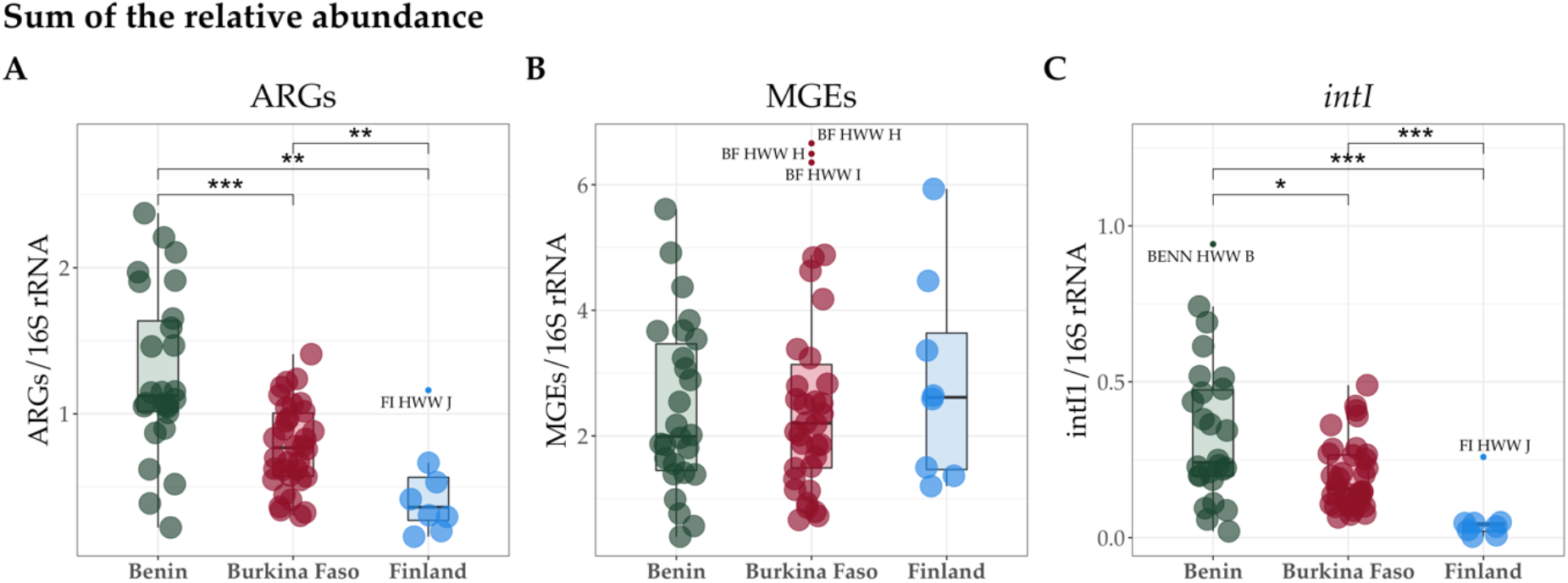
The sum of the relative abundance of A) ARGs, B) all MGEs, and C) class 1 integrons (*intiI1*). The gene counts were normalized to 16S rRNA gene counts and gene lengths. Country medians are shown as vertical lines and the interquartile ranges (25^th^ and 75^th^ percentiles) as boxplot hinges. The horizontal lines represent the highest and lowest values. Outliers are defined as values higher or lower than 1.5 times the upper or lower percentiles, respectively, and denoted with a text label referring to the category represented by that sample. The comparisons between countries were computed using pair-wise Wilcoxon rank sum tests adjusted by Benjamini-Hochberg. The significance levels are ‘*’ = ≤ 0.05; ‘**’ ≤ 0.01; ‘***’ = ≤ 0.001.

According to the Kruskal-Wallis test, there were significant (*p*-value < 0.005) country-wise differences in the sums of relative abundances of ARGs and *intI1*, but not when all MGEs were included (Figure 1B). The significant differences were investigated further using Wilcoxon rank sum tests adjusted by Benjamini-Hochberg. The differences in class 1 integron genes (*intI1*) followed a similar pattern in country-wise comparisons as ARGs (Figure 1C). In fact, the correlations between the counts of ARGs and class 1 integron genes, *intI1* and *qacEΔ*, were stronger for HWW samples from Benin compared to Burkina Faso and Finland (Figure S4A, S4B). Additionally, contigs with multiple ARGs, such as carbapenemase or ESBL variants of *bla*GES and quinolone resistance genes (*qnrVC*) located in proximity to each other, were identified (Figure S5). These contigs might indicate gene cassettes carried by class 1 integron elements as previously reported for these genes (*bla*GES and *qnrVC*) in various ARG combinations ^10,35,36^. In contrast to *intI1*, no significant correlations were observed between ARGs and *int2* or *int3* with HWW from Benin (Figure S4C, S4D), unlike with HWW from Burkina Faso (Figure S4C, S4D).

Compositional data analysis (CoDa) methods were applied to investigate the ordination of the samples from the different countries by their resistome, taxonomical composition, and mobilome with respect to each other. HWW resistomes from Benin and Burkina Faso resembled each other and formed clusters clearly distinct from the resistomes from Finland. (Figure 2A). Country explained less of the variance between taxonomical compositions than between resistomes (Figure 2B), while similarly to resistomes, distinct country-wise clusters were seen with the mobilomes (Figure 2C). Eleven non-HWW water samples, such as those collected from rivers and near or within the hospital environment, were analyzed to obtain a more comprehensive understanding of the ARG prevalence in Benin and Burkina Faso. These samples were located within the clusters of their respective countries when ordinated together with the HWW (Figure S6). Since the HWW resistomes from Benin and Burkina Faso differed notably from the resistomes from Finland, the drivers for these differences were subsequently investigated.

**Figure 2.**
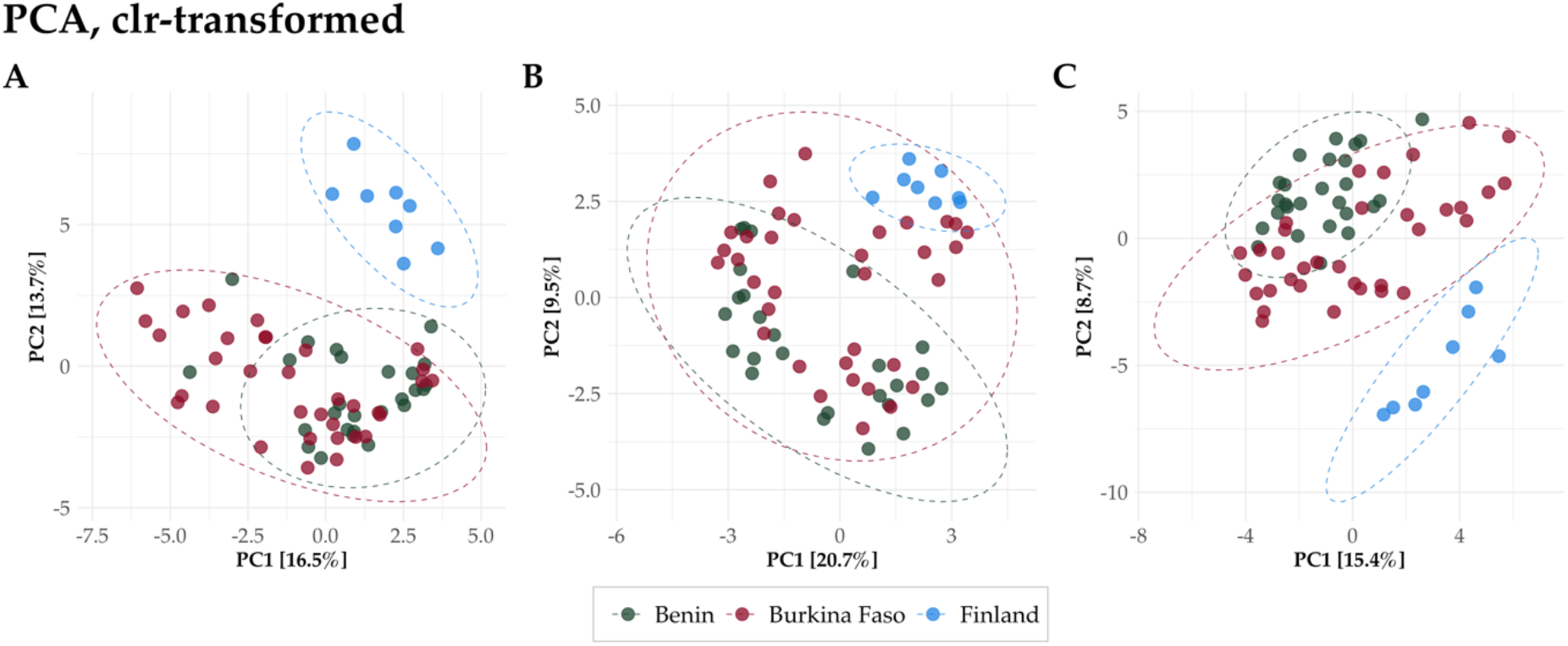
Principal Components Analysis (PCA) showing the significant dissimilarities of A) resistomes, B) taxonomical composition Metaphlan3, and C) mobilomes in HWW from Benin, Burkina Faso, and Finland. Count data were clr-transformed. Confidence ellipses represent 95 % confidence levels.

### 2.2. Significantly more abundant ARGs

ARGs significantly more abundant in HWW from each country-wise comparison were investigated. These differentially abundant ARGs in the HWW resistomes from Benin, Burkina Faso, and Finland were revealed by applying compositional analysis by ANOVA-Like Differential Expression tool for high throughput sequencing data (ALDEx2) ^37^ tool. In this analysis, 16S rRNA gene counts were used as the denominator for the alr-transformation of the count data.

In comparisons Benin – Finland and Burkina Faso – Finland, ARGs significantly more abundant in HWW from Finland were fewer in number than those from Benin or Burkina Faso (Figures 3A and 3B) (BENN (n = 258) – FI (n = 33); BF (n = 257) – FI (n = 29)). In addition, the ARGs that were significantly more abundant in HWW from Finland in both comparisons (Benin – Finland and Burkina Faso – Finland) were primarily the same (Tables 2, S3, and S4). *bla*OXA-211-like cluster genes and *erm(B)* macrolide resistance genes were ARGs significantly more abundant in HWW from Finland, as seen in the list in Table 2 of the top 10 significantly more abundant ARGs by their ‘diff.btw’ values, which represent the median difference in the alr-transformed count data between the compared HWW (see Tables S3-S5 for all results). The ARGs characteristic for HWW from Benin included quinolone resistance gene *qnrVC*, ESBL genes *bla*VEB, *bla*CARB, and *bla*OXA-5-like cluster genes (Tables 2 and S3). For Burkina Faso, significantly more abundant ARGs were *bla*CMY genes, trimethoprim resistance genes of *dfrA15* and *dfrB* clusters, and genes of *bla*OXA-10- and *bla*OXA-427-like clusters (Tables 2 and S4).

**Table 2.** Top 10 differentially abundant ARGs in HWW from Benin, Burkina Faso, and Finland. The ‘diff.btw’ values represent the median difference in the alr-transformed count data between the compared HWW. The expected Benjamini-Hochberg corrected *p*-values of the Wilcoxon test are indicated in the table as ‘wi.eBH’. The ARG clustering defined the cluster names using CD-HIT ^39^ based on 90 % of shared sequence identity.

| Benin - Finland |  |  |  |  |
| --- | --- | --- | --- | --- |
| diff.btw | wi.eBH | gene | cluster name | country |
| -8.5066 | 5.78E-05 | *lnu(F)_3* | *lnu(F)* | Benin |
| -7.7019 | 4.32E-04 | *aac(6')-IIc_1_NC* | *aac(6')-IIc* | Benin |
| -7.4755 | 7.69E-05 | *qnrVC5_1* | *qnrVC4* | Benin |
| -7.4752 | 3.74E-05 | *qnrVC4_1* | *qnrVC4* | Benin |
| -7.2563 | 1.33E-04 | *blaCARB-2_1* | *blaCARB-50* | Benin |
| -6.8960 | 1.74E-04 | *ant(2'')-Ia_6* | *ant(2'')-Ia_2* | Benin |
| -6.7910 | 6.06E-03 | *aph(2'')-Id_1* | *aph(2'')-Id_1* | Benin |
| -6.7897 | 7.05E-05 | *blaVEB-5_1* | *blaVEB-1* | Benin |
| -6.6886 | 4.20E-05 | *blaVEB-2_1* | *blaVEB-1* | Benin |
| -6.6295 | 4.69E-05 | *blaVEB-1_1* | *blaVEB-1* | Benin |
| 4.7351 | 1.13E-04 | *blaOXA-309_1* | *blaOXA-211-like* | Finland |
| 4.7739 | 4.40E-04 | *blaOXA-427_1* | *blaOXA-427-like* | Finland |
| 4.8342 | 8.51E-05 | *erm(B)_21* | *erm(B)_18* | Finland |
| 5.0383 | 5.46E-04 | *blaOXA-280_1* | *blaOXA-211-like* | Finland |
| 5.3202 | 1.18E-04 | *erm(B)_10* | *erm(B)_18* | Finland |
| 5.3388 | 2.25E-04 | *blaOXA-334_1* | *blaOXA-211-like* | Finland |
| 5.4158 | 6.85E-03 | *blaOXA-333_1* | *blaOXA-211-like* | Finland |
| 5.4932 | 7.92E-05 | *blaOXA-212_1* | *blaOXA-211-like* | Finland |
| 5.9308 | 6.52E-05 | *blaOXA-373_1* | *blaOXA-211-like* | Finland |
| 6.5027 | 2.08E-03 | *blaOXA-211_1* | *blaOXA-211-like* | Finland |

| Burkina Faso - Finland |  |  |  |  |
| --- | --- | --- | --- | --- |
| diff.btw | wi.eBH | gene | cluster name | country |
| -7.1539 | 1.59E-05 | *blaCMY-4_1* | *blaCMY-150* | Burkina Faso |
| -6.9233 | 3.12E-05 | *blaOXA-101_1* | *blaOXA-10* | Burkina Faso |
| -6.1860 | 1.76E-05 | *dfrA15_2* | *dfrA15* | Burkina Faso |
| -6.1384 | 3.10E-04 | *dfrB5_1* | *dfrB1* | Burkina Faso |
| -5.9944 | 7.31E-05 | *dfrA15_1* | *dfrA15* | Burkina Faso |
| -5.9796 | 3.12E-04 | *blaCMY-22_1* | *blaCMY-150* | Burkina Faso |
| -5.9309 | 6.85E-05 | *blaCMY-121_1* | *blaCMY-150* | Burkina Faso |
| -5.9133 | 1.02E-04 | *blaCMY-32_1* | *blaCMY-150* | Burkina Faso |
| -5.8866 | 6.73E-04 | *blaCMY-133_1* | *blaCMY-150* | Burkina Faso |
| -5.8772 | 1.40E-04 | *blaCMY-69_1* | *blaCMY-150* | Burkina Faso |
| 4.7242 | 2.79E-03 | *erm(B)_21* | *erm(B)_18* | Finland |
| 5.2610 | 5.69E-05 | *blaOXA-309_1* | *blaOXA-211-like* | Finland |
| 5.2630 | 4.30E-05 | *blaOXA-334_1* | *blaOXA-211-like* | Finland |
| 5.3117 | 2.68E-04 | *blaOXA-281_1* | *blaOXA-211-like* | Finland |
| 5.3789 | 2.29E-04 | *blaOXA-373_1* | *blaOXA-211-like* | Finland |
| 5.4740 | 5.13E-04 | *blaOXA-280_1* | *blaOXA-211-like* | Finland |
| 5.4817 | 5.47E-05 | *blaOXA-299_1* | *blaOXA-211-like* | Finland |
| 5.7319 | 3.83E-03 | *blaOXA-333_1* | *blaOXA-211-like* | Finland |
| 6.0623 | 7.13E-05 | *blaOXA-212_1* | *blaOXA-211-like* | Finland |
| 7.1596 | 1.58E-03 | *blaOXA-211_1* | *blaOXA-211-like* | Finland |

| Benin - Burkina Faso |  |  |  |  |
| --- | --- | --- | --- | --- |
| diff.btw | wi.eBH | gene | cluster name | country |
| -5.5058 | 2.15E-08 | *blaOXA-129_1* | *blaOXA-5* | Benin |
| -5.3352 | 1.65E-06 | *aac(6')-IIc_1_NC* | *aac(6')-IIC* | Benin |
| -5.1830 | 3.49E-05 | *aph(2'')-Ib_1* | *aph(2'')-Ib_2* | Benin |
| -5.1784 | 3.87E-05 | *aac(6')-Im_1* | *aac(6')-Im* | Benin |
| -4.2138 | 4.40E-04 | *aph(2'')-Ib_2* | *aph(2'')-Ib_2* | Benin |
| -3.8822 | 6.43E-04 | *sul3_2* | *sul3* | Benin |
| -3.7533 | 2.56E-03 | *aph(2'')-Ib_3* | *aph(2'')-Ib_2* | Benin |
| -3.5422 | 1.89E-02 | *aph(2'')-Id_1* | *aph(2'')-Id* | Benin |
| -3.4469 | 1.63E-04 | *lnu(C)_1* | *lnu(C)* | Benin |
| -3.3127 | 7.05E-04 | *lnu(F)_3* | *lnu(F)* | Benin |
| 2.6181 | 2.22E-02 | *blaOXA-420_1* | *blaOXA-58-like* | Burkina Faso |
| 2.7238 | 1.75E-02 | *blaOXA-96_1* | *blaOXA-58-like* | Burkina Faso |
| 2.8240 | 1.32E-02 | *blaOXA-397_1* | *blaOXA-58-like* | Burkina Faso |
| 2.8658 | 1.58E-02 | *blaOXA-97_1* | *blaOXA-58-like* | Burkina Faso |
| 2.8783 | 1.82E-02 | *blaOXA-164_1* | *blaOXA-58-like* | Burkina Faso |
| 2.9017 | 8.67E-04 | *tet(39)_1* | *tet(39)* | Burkina Faso |
| 3.1943 | 2.06E-02 | *blaBEL-2_1* | *blaBEL-1* | Burkina Faso |
| 3.4107 | 2.88E-03 | *blaOXA-58_1* | *blaOXA-58-like* | Burkina Faso |
| 3.5374 | 1.04E-02 | *blaBEL-3_1* | *blaBEL-1* | Burkina Faso |
| 4.0895 | 2.50E-03 | *blaBEL-1_1* | *blaBEL-1* | Burkina Faso |

**Figure 3.**
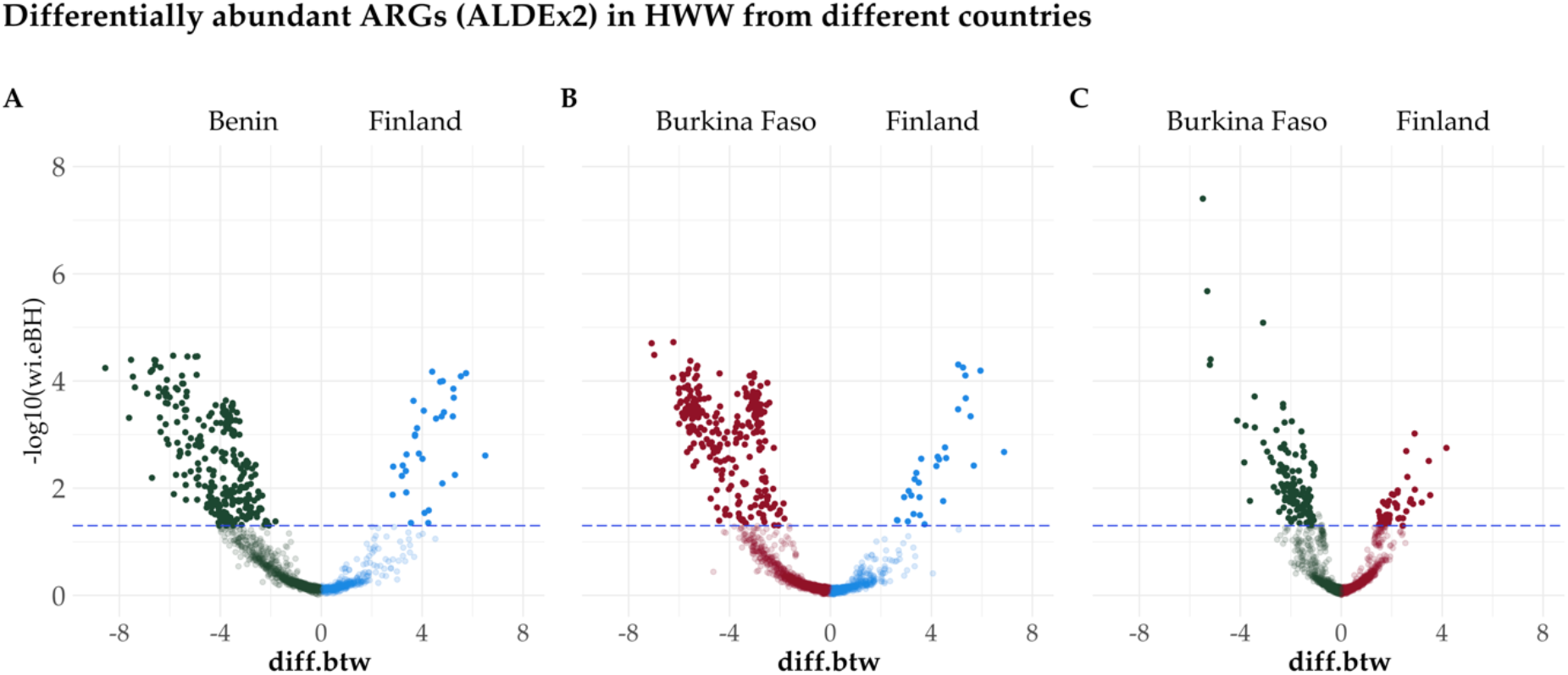
Significantly more abundant ARGs in HWW in pair-wise comparisons in Benin – Finland, Burkina Faso – Finland, and Benin – Burkina Faso were defined using the ALDEx2 tool ^38^ with 16S rRNA as the reference gene. For instance, in Figure 3A, significantly more abundant ARGs for Benin (green dots above the blue dotted line) have negative ‘diff.btw’ values (x-axis), while ARGs significantly more abundant for Finland (blue dots above the blue dotted line) have positive values. The dotted line represents a *p* < 0.05 of the expected Benjamini-Hochberg corrected *p*-value of the Wilcoxon test (wi.eBH). These *p*-values are log10-transformed for the figure.

The comparison between HWW from Benin and Burkina Faso revealed fewer differentially abundant ARGs between these countries (BENN (n = 49) – BF (n = 119)) than in the comparisons with Finnish HWW. In addition, the volcano in Figure 3C is more skewed towards the vertical blue dotted line, thus referring to less drastic differences in the abundances of the ARGs in this comparison. These results support the previous notion that the resistomes in HWW from Benin and Burkina Faso were more similar to each other than those from Finland (Table 2, Tables S3-S5). However, ESBL genes of *bla*BEL, and carbapenemases of the *bla*OXA-58-like cluster, were significantly more abundant in HWW from Burkina Faso than from Benin (Tables 2 and S5). Instead, different aminoglycoside resistance genes and lincosamide resistance genes *lnu(F)* and *lnu(C)* were characteristic for Benin (Tables 2 and S5).

As a further notion, those *bla*OXA variants that were significantly more abundant in HWW from Finland than the ones in Benin and Burkina Faso were predominantly those that are intrinsically carried by some specific species and encoding carbapenemases (e.g., *bla*OXA-211-like genes) (Table 3). Instead, the *bla*OXA variants characteristic for HWW from Benin and Burkina Faso were those that are typically acquired and, further, do not encode carbapenemase activity (e.g., *bla*OXA-5 and *bla*OXA-10) (Table 3). For example, *bla*OXA-5-like clusters, significantly more abundant in Benin versus Burkina Faso or Finland, were those typically carried by class 1 integrons ^40^.

**Table 3.**
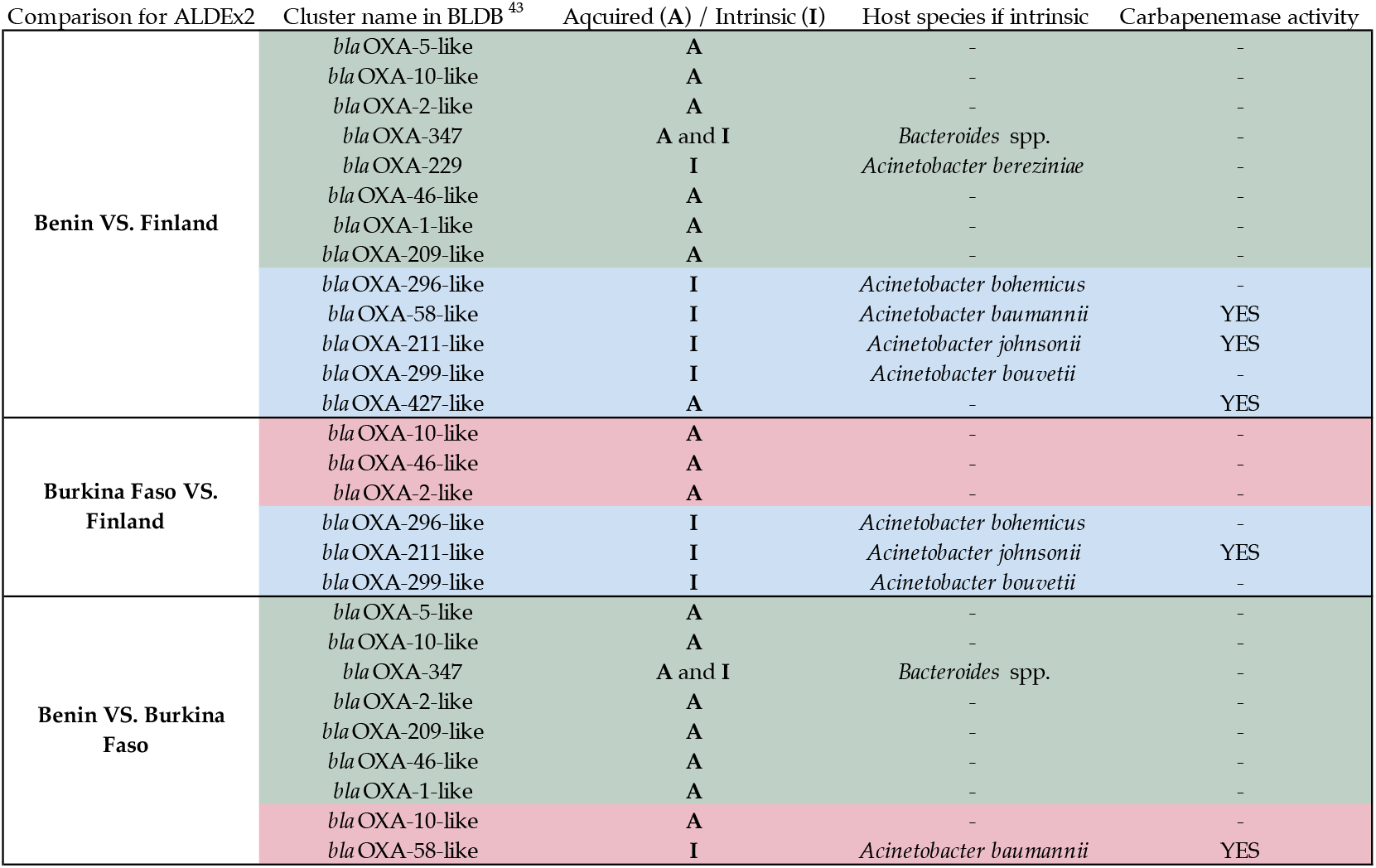
The differentially abundant *bla*OXA variants in HWW from Benin, Burkina Faso, and Finland. The ARGs significantly more abundant in each country are denoted with color encoding; Benin = green, Burkina Faso = red, Finland = blue. The data concerning the origin of the *bla*OXA gene, whether it is intrinsic or acquired, was retrieved from the data collected to Beta Lactamase DataBase (BLDB) ^41^, except that we used the expression ‘intrinsic’ instead of the ‘natural in the original text, due to the misleading connotation of the word ‘natural’ in the context of antibiotic resistance.

### 2.3. Carbapenemases

The presence of seven acquired carbapenemases (*bla*GES, *bla*IMP, *bla*KPC, *bla*NDM, *bla*OXA-48, *bla*OXA-58, and *bla*VIM) was analyzed in detail due to their ability to cause complex infections with minimal treatment options especially in LMICs when in a relevant host. The highest relative abundance of these carbapenemases was observed in the Finnish hospital J (Figure 4). On the other hand, hospital M in Finland was nearly free from these carbapenemases (Figure 4). In HWW from Benin, the carbapenemase genes in the *bla*GES gene family seemed to dominate over other carbapenemases in all four hospitals (Figure 4). *bla*GES genes were also detected in the water puddle surrounding the surgery room septic tank at a Beninese hospital (hospital C) yard (Figure 4, Figure S2E) as well as in the water intended for hand-washing in the same hospital (hospital C) (Figure 4, Figure S2F). Also, the street gutter, located approximately 100 m away from another Beninese hospital, was contaminated by *bla*GES carbapenemases (Figure 4).

**Figure 4.**
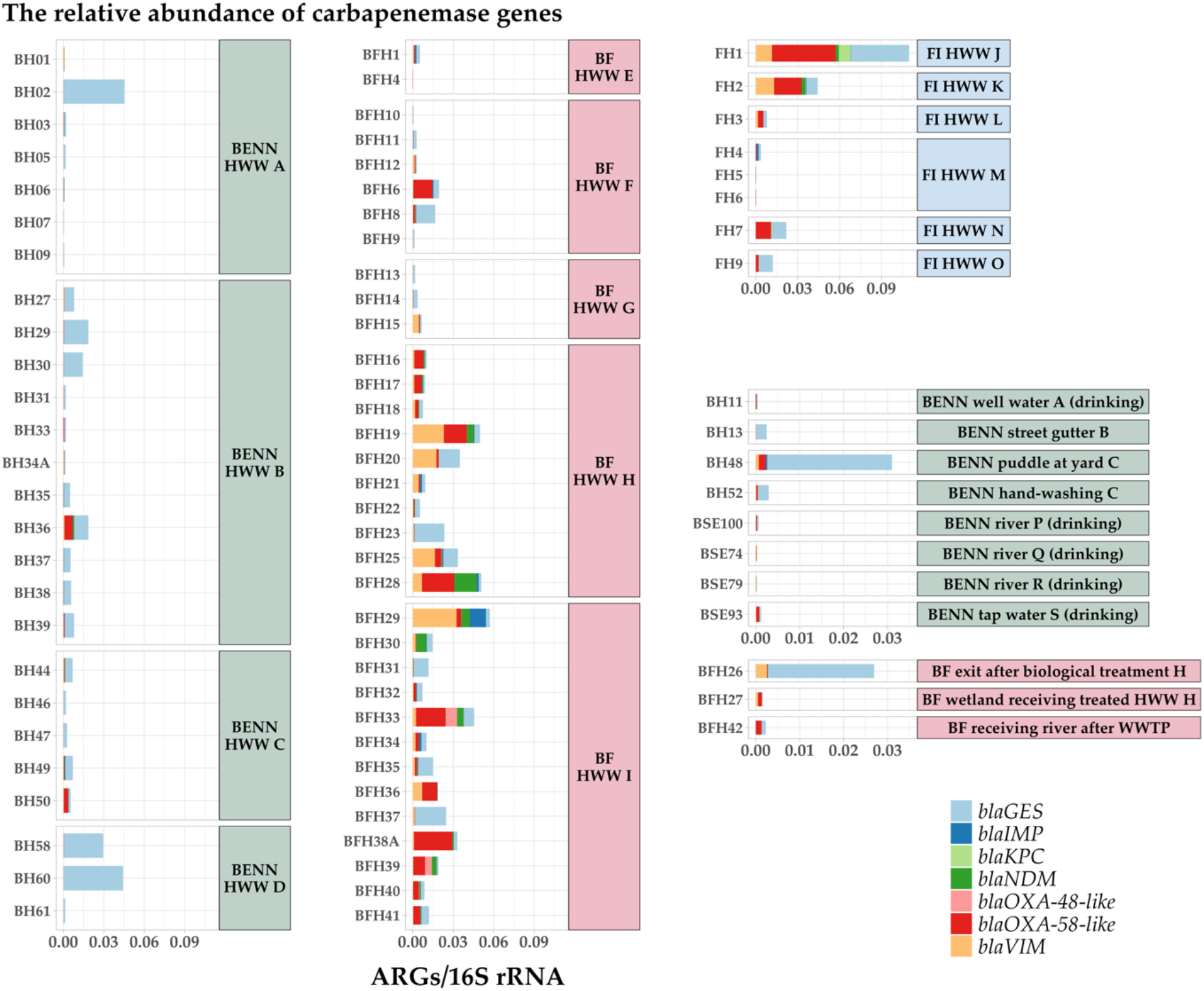
The relative abundance of carbapenemase genes in relation to 16S rRNA gene counts in HWW from Benin (left, green label background), Burkina Faso (center, red label background), and Finland (up-right, blue label background), and various other water sources in Benin (center-right, green label background) and Burkina Faso (bottom-right, red label background). Note the differences in the y-axis scales between the figures for HWW and non-HWW samples. Only variants known to encode carbapenemases were screened ^41^ (Table S6).

In contrast to the homogeneity of carbapenemases in HWW from Benin, most of the other carbapenemases were present in HWW from Burkina Faso and Finland at various prevalence levels (Figure 4). *bla*IMP, *bla*NDM, and *bla*VIM were mainly detected in Burkinabe and Finnish HWW samples, and the latter was significantly more abundant in HWW from Burkina Faso (Figure 4, Table S4). *bla*OXA-48 was detected in two HWW samples from Burkina Faso and not at all or in very low relative abundances in samples from elsewhere (Figure 4). Instead, *bla*OXA-58-like genes were present in the majority of Burkinabe and Finnish HWW samples but only in a few Beninese samples (Figure 4). The detection of *bla*KPC genes was restricted to one Finnish HWW sample (Figure 4). The samples collected from natural waters and other drinking waters showed only low relative abundances of these carbapenemases (Figure 4). However, some *bla*OXA-58-like were detected in tap water used for drinking in Benin (Figure 4).

In Burkina Faso, *bla*GES carbapenemases were detected in HWW, which had gone through the biological treatment in similar relative abundances as in some of the samples of untreated HWW from the same hospital (hospital H) (Figure 4, note the differences in the plot scales). Also, *bla*VIM was found both in the untreated and treated water of that hospital (Figure 4). Lower relative abundances of the studied carbapenemases were observed in wetlands and rivers receiving treated HWW in Burkina Faso (Figure 4). However, the spectrum of these carbapenemases, namely *bla*OXA-58-like, *bla*VIM, and *bla*GES, reflected the ones detected in the HWW in Burkina Faso (Figure 4).

### 2.4. Mobile colistin resistance (*mcr*) genes

*mcr* genes were detected in several HWW samples in Benin, Burkina Faso, and Finland (Table 4). *mcr-5* (variants *mcr-5*.*1* and *mcr-5*.*2*, Table S7) was the most common of the *mcr* genes as they were found in HWW from all except one hospital (hospital E) among the three countries. In the few samples selected for metagenomic assembly, this gene was found to be located within a Tn3-like element (Figure S7).

**Table 4.** The relative abundances of *mcr* genes. For the HWW samples, averages for relative abundances (average RA) and the number of positive samples (n) are shown.

|  |  | BENN HWW | BF HWW | FI HWW | BENN well water A<br>(drinking) (BH11) | BENN street<br>gutter B (BH13) | BENN puddle<br>at yard C<br>(BH48) | BENN hand-washing C (BH52) |
| --- | --- | --- | --- | --- | --- | --- | --- | --- |
| *mcr-1* | average RA<br>n | 0<br>0/26 | 4.61E-07<br>1/34 | 1.97E-06<br>1/8 | 0 | 0 | 0 | 0 |
| *mcr-2* | average RA<br>n | 0<br>0/26 | 0<br>0/34 | 0<br>0/8 | 0 | 0 | 0 | 0 |
| *mcr-3.1* | average RA<br>n | 6.41E-05<br>16/26 | 4.18E-04<br>18/34 | 1.58E-04<br>7/8 | 4.24E-05 | 9.71E-03 | 1.96E-02 | 1.77E-03 |
| *mcr-3.17* | average RA<br>n | 2.26E-06<br>2/26 | 3.74E-06<br>9/34 | 1.32E-04<br>5/8 | 0 | 7.76E-05 | 1.23E-04 | 0 |
| *mcr-4* | average RA<br>n | 3.42E-06<br>1/26 | 2.63E-06<br>3/34 | 0<br>0/8 | 0 | 0 | 0 | 0 |
| *mcr-5* | average RA<br>n | 8.92E-04<br>24/26 | 1.02E-03<br>21/34 | 7.53E-04<br>8/8 | 4.19E-05 | 2.25E-03 | 1.99E-03 | 7.76E-05 |
| *mcr-6* | average RA<br>n | 0<br>0/26 | 0<br>0/34 | 0<br>0/8 | 0 | 0 | 0 | 0 |
| *mcr-7* | average RA<br>n | 5.06E-05<br>10/26 | 1.54E-04<br>25/34 | 1.94E-04<br>8/8 | 0 | 1.09E-04 | 7.07E-05 | 3.94E-05 |
| *mcr-8* | average RA<br>n | 0<br>0/26 | 0<br>0/34 | 1.68E-06<br>1/8 | 0 | 0 | 0 | 0 |
| *mcr-9* | average RA<br>n | 1.04E-06<br>1/26 | 2.14E-06<br>7/34 | 3.08E-04<br>4/8 | 0 | 3.11E-05 | 0 | 0 |
| *mcr-10* | average RA<br>n | 1.33E-04<br>12/26 | 1.03E-03<br>20/34 | 1.97E-05<br>2/8 | 4.25E-05 | 1.24E-04 | 1.77E-05 | 3.94E-05 |

|  |  | BENN tap<br>water S<br>(drinking)<br>(BSE93) | BENN<br>river P<br>(drinking)<br>(BSE100) | BENN<br>river Q<br>(drinking)<br>(BSE74) | BENN river R<br>(drinking) (BSE79) | BF exit after<br>biological<br>treatment H<br>(BFH26) | BF wetland<br>receiving<br>treated HWW H<br>(BFH27) | BF receiving river<br>after WWTP<br>(BFH42) |
| --- | --- | --- | --- | --- | --- | --- | --- | --- |
| *mcr-1* | average RA<br>n | 0 | 0 | 0 | 0 | 0 | 0 | 0 |
| *mcr-2* | average RA<br>n | 0 | 0 | 0 | 0 | 0 | 0 | 0 |
| *mcr-3.1* | average RA<br>n | 2.43E-05 | 0 | 0 | 0 | 4.19E-05 | 8.70E-05 | 2.65E-05 |
| *mcr-3.17* | average RA<br>n | 0 | 3.23E-05 | 0 | 0 | 0 | 0 | 0 |
| *mcr-4* | average RA<br>n | 0 | 0 | 0 | 0 | 0 | 0 | 0 |
| *mcr-5* | average RA<br>n | 0 | 0 | 0 | 1.10E-04 | 2.80E-02 | 4.30E-04 | 2.36E-04 |
| *mcr-6* | average RA<br>n | 0 | 0 | 0 | 0 | 0 | 0 | 0 |
| *mcr-7* | average RA<br>n | 9.74E-05 | 0 | 0 | 3.73E-05 | 8.40E-05 | 0 | 0 |
| *mcr-8* | average RA<br>n | 0 | 0 | 0 | 0 | 0 | 0 | 0 |
| *mcr-9* | average RA<br>n | 0 | 3.23E-05 | 0 | 0 | 0 | 0 | 0 |
| *mcr-10* | average RA<br>n | 4.87E-05 | 0 | 0 | 0 | 0 | 1.74E-04 | 7.97E-05 |

In addition to the HWW samples, high relative abundances of *mcr-5* genes were also detected in the immediate and more distant surroundings of the hospitals in Benin and Burkina Faso. A very high relative abundance (2.80 × 10^−2^) of *mcr-5* was observed in the biologically treated HWW from hospital H (Table 4). In the wetland receiving treated HWW from the same hospital, the relative abundance was lower (4.30 × 10^−4^) but still high, considering that the sample represented a larger body of natural water. In Benin, *mcr-5* was detected in river water in a distant village and a street gutter near a hospital (hospital B) (Table 4). Even the water intended for hand-washing in a Beninese hospital (hospital C) was not free from *mcr-5*. Furthermore, for hospital C, the relative abundance of *mcr-5* was greater in a puddle near the HWW containing septic tanks in the hospital yard (1.99 × 10^−3^, Table 4) than in the actual HWW of that hospital (5.24 × 10^−4^, data not shown).

The next most prevalent *mcr* genes were *mcr-3, mcr-10*, and *mcr-7*, and similarly to *mcr-5*, they were present also in non-HWW samples (Table 4). The gene *mcr-2* was not detected at all, and the lowest average relative abundance was detected for the gene *mcr-1* (Table 4).

### 2.5. Taxonomical compositions

The taxonomical composition of bacteria present in septic tanks and sumps in Benin and Burkina Faso differed from those found in hospital sewers in Finland. Genera belonging to the *Bacteroidales* family had a significantly higher abundance in HWW from Benin and Burkina Faso compared to Finland (Figures S8 and S9). Other significantly more abundant taxa in HWW from Benin compared to Finland included *Chloracidobacterium, Geobacter, Aminomonas, Flexilinea, Desulfovibrio*, and genera of *Synergistetes* (Figure S8A, Table S8). The first two mentioned were characteristic also for the HWW from Burkina Faso (Figure S8B, Table S9). In addition, in many HWW samples from Burkina Faso, the relative abundances of *Pseudomonas* and *Acinetobacter* were high (Figure S9), and the difference in their abundance in HWW from Burkina Faso versus Benin was significant (Figure S8C, Table S10).

*Coprococcus, Enterococcus, Lactococcus, Streptococcus, Trichococcus, Tessaracoccus, Delftia, Raultella*, and genera of the *Proteobacteria* family, were all significantly more abundant in HWW from Finland in comparison to Benin and Burkina Faso (Figures S8A and S8B, Tables S8 and S9). Also, the genera that were significantly more abundant for HWW in Finland and not Benin were more commonly from the phyla (*Pseudomonadota*, synonym *Proteobacteria*), which are typically considered as the major contributors to the spread of ARGs carried by plasmids, integrons, or other MGEs ^10,42^ (Figure S8).

A great variation was seen in the top 11 taxa in the non-HWW samples (Figure S9). The biologically treated HWW from Burkina Faso showed high relative abundances of *Aeromonas* and *Pseudomonas*. In contrast, high relative abundances of *Bacteroides* and *Klebsiella* were detected in the street gutter water located near hospital B (Figure S9). In some samples, the top 11 taxa were present only in low relative abundances, while in contrast, a single genus of *Polynucleobacter* dominated over other genera in a few sampled natural waters receiving treated wastewater, both in Benin and Burkina Faso (Figure S9). The relative abundance of *Acinetobacter* in tap water used for drinking in Benin reached a similar level that in many HWW samples (Figure S9), which possibly explained the finding of *bla*OXA-58-like genes in this sample mentioned earlier (Figure 3).

## 3. Discussion

We characterized the bacterial community composition, resistome, and mobilome of 60 HWW and 11 other water samples from Benin and Burkina Faso and compared them to 8 HWW samples from Finland. Due to the lack of systematic AMR surveillance in these West African countries, available data on bacterial resistance are patchy and heterogeneous 1,4,23,26,29. Thus, the magnitude of the resistance problem and the specific ARG reservoirs are yet to be unraveled. This is the first study investigating hospital wastewater in Benin and Burkina Faso using a shotgun metagenomic approach. Both ‘traditional’ normalization methods, and compositional data analysis ^43^, were applied in the analyses. For the latter, the ALDEx2 tool was chosen over Differential gene expression analysis based on the negative binomial distribution (DESeq2) ^44^ as we discovered that the significantly differentially abundant ARGs were reported less strictly with DESeq2 than ALDEx2, and we wanted to avoid possible false positive results generated by DESeq2 (Figure S11C).

Interestingly, the ARGs observed in HWW from Benin were the highest in number but probably less clinically important than those in the HWW from Burkina Faso and Finland. While carbapenemases *bla*GES, *bla*IMP, *bla*NDM, *bla*OXA-48-like, *bla*OXA-58-like, and *bla*VIM were detected at various abundances in at least one sample in HWW from Burkina Faso and Finland, *bla*GES was the predominant carbapenemase in HWW from Benin. *bla*GES genes were also present in the water puddle at the Beninese hospital’s yard and in water intended for visitors’ hand-washing in the hospital. Thus, one route of transmission of these ARGs within hospitals might be via hands contaminated by hand-washing water, which might explain the high prevalence of these genes in the hospitals. Jacobs and colleagues have also drawn attention to the potentially inferior microbiological water quality in similar hand-washing water tanks, which are very common in West Africa ^4^.

Furthermore, we speculate that the dominance of *bla*GES carbapenemases might have been caused by a selection pressure due to the presence of antibiotic residues in the HWW. However, as carbapenem antibiotics are used less in West African countries than in North America and Southern and Central Europe, due to their high price ^16^, we suggest that compounds from other antibiotic classes could have driven the selection. In fact, *bla*GES genes are typically carried by class 1 integrons in which they may be coupled with multiple other ARGs ^45^. Thus, we speculate that class 1 integrons ^14^ and co-selection phenomenon ^3,46^ could have a role in the dominance of *bla*GES. That is, in the example of metagenome-assembled contigs in which quinolone ARGs (*qnrVC*) and *bla*GES genes seemed to be located near each other putatively encoded by the class 1 integron gene cassette, the selection pressure targeted to the quinolone ARG would enrich both genes even in the absence of the target substrate for *bla*GES. As a potential limitation of our study, we agree that there is a high sequence similarity among *bla*GES variants, including those conferring ESBL and carbapenem resistance. Therefore, we acknowledge the possibility that the putative lack of specificity related to the read mapping might have resulted in some misidentifications among the different variants of this gene.

Despite the lowest sum of the relative abundance of ARGs detected in HWW from Finland, some of the seven carbapenemase genes showed higher relative abundances in Finland than in HWW of the surveyed West African hospitals. For the occurrence of *bla*OXA genes, the abundances of specific species present in the different HWW collection systems might have played a role, as *bla*OXA variants that encode carbapenemases and are typically intrinsically carried by certain *Acinetobacter* species ^47,48^ were significantly more abundant in Finnish HWW. Instead, those *bla*OXA variants that are typically acquired and mainly encode more narrow spectrum beta-lactamases were significantly more abundant in HWW from Benin and also Burkina Faso ^41^. Although, the occurrence of the seven carbapenemases was not homogenous among the Finnish samples. For example, *bla*KPC was restricted to a single HWW sample from Finland (hospital J) and none from Benin or Burkina Faso. This finding aligns with local and global reports and systematic reviews ^49–52^, indicating that *bla*KPC carbapenemases are spreading more profusely in Europe and North America than in Africa. Moreover, the two hospitals, J and K, both part of the University Central Hospital of the capital of Finland, showed the highest sum of relative abundance of the seven carbapenemase genes among all HWW samples in this study. These results are somewhat surprising as, to date, Finland is known as one of the countries with the lowest level of bacterial resistance in Europe and globally ^53^ and the carbapenemase-producing strains detected in Finnish hospitals are relatively rare and typically associated with international travel or hospitalization ^54^.

One factor likely explaining our findings of the distinctive features in the HWW resistomes in Benin and Burkina Faso compared to Finland was the differences in the wastewater collection systems between the studied countries. In Finland, the hospital toilet waters containing human fecal material are directed to HWW, while for the majority of the Burkinabe and especially Beninese HWW studied here, this was not the case. Additionally, in septic tanks (Benin and Burkina Faso), the water remains stagnant, possibly giving rise to the anaerobes, unlike the Finnish HWW, which flows through the system. Significant prevalence of anaerobic genera such as *Geobacter, Aminomonas, Flexilinea, Desulfovibrio*, and genera of *Synergistetes* and *Bacteroidales* observed in HWW from Benin and Burkina Faso and not in Finland is in line with this speculation except for the last-mentioned taxa, which is a typical human gut commensal ^55^. These findings also align with the previous reports on bacterial genera typically found in soil and aquatic environments ^56^ and previously described to dominate HWW from Benin and Burkina Faso ^57^. Facultatively anaerobic bacteria such as the genera of lactic acid bacteria, *Aeromonas*, and the anaerobic *Bifidobacterium* ^58^ which are considered typical human gut microbes ^55^ were significantly more abundant in the Finnish HWW than in Benin or Burkina Faso.

Compared to many high-income countries such as those in Northern Europe, antibiotic usage in agricultural ^6,7^ and clinical ^4^ settings differ ^16^ and is less controlled – or even unregulated – in many African countries. For example, while banned in many other countries, the use of colistin as a feed additive is allowed in many LMICs ^59^, also in Africa ^7^. *mcr-5* was the most commonly detected *mcr*-gene in the HWW in our study, contrary to previous reports on the prevalence of *mcr* genes globally ^22^ and in Africa ^23,60^. This may be due to the methodologies used to screen for colistin resistance. Other than the best-known *mcr*-genes (*mcr-1, -2*, and *-3*) are rarely targeted when screening for *mcr* genes using conventional PCR ^61^. Therefore, our study shows that to obtain a more realistic view of *mcr* genes in Africa, screening should be conducted for a broader set of different *mcr* genes, as was recently done by Ngbede and colleagues ^62^.

The *mcr-5* gene detected was embedded in a Tn3-like element similar to previous reports for *Salmonella enterica* ^63,64^ and *Escherichia coli* ^65^ plasmids and the chromosome of *Cupriavidus gilardii* ^63^. These Tn3-like elements are flanked by inverted repeats, which enable translocation and putatively a broad host range for the *mcr-5* harboring element ^63–65^. Based on our results, the occurrences of *mcr-5* and other *mcr* genes in Benin and Burkina Faso were not restricted to HWW septic tanks. These genes were also detected in water intended for the hand-washing of visitors to the hospital. The high prevalence of *mcr-5* among our various samples here raises questions about its origin and ecology, whether it is of clinical origin or intrinsically carried by some environmental bacteria. However, the highest relative abundances of *mcr-5* were observed in samples influenced by human activity, and three of the four rivers in the remote village were free from this gene.

In Burkina Faso, the relative abundance of *mcr-5* in HWW, even after the biological treatment, was high. Although we could not confirm the association between the *mcr-5* genes detected in the treated and the hospital-associated wastewater in the Burkinabe hospital, our study suggests the inability of the currently used wastewater treatment processes in Burkina Faso to remove the *mcr* genes. Wastewater treatment systems have indeed previously been described as inadequate in Benin and Burkina Faso ^66^. The situation appears to be especially critical in Cotonou, where the hospitals involved in the study did not use any kind of wastewater treatment. Groundwater in the city is extracted from the shallow aquifer, which is polluted due to unauthorized waste deposits, inadequate toilets, pit latrines, and septic tanks prone to leakage and hydraulic failure ^67^. This enables the continuous circling of *mcr* genes and other ARGs as the inadequately treated water is released into natural waters and used for various purposes by local people. Furthermore, in Africa, untreated wastewater is commonly used for irrigation in urban agriculture, possibly enabling the dissemination of ARGs to fresh produce ^68,69^. Our study shows that untreated or even treated hospital wastewater, which can leak into groundwater and the environment, may carry clinically hazardous antibiotic-resistant bacteria or resistance genes.

Taken together, based on our results from nine hospitals, we can state that clinically important ARGs are circulating in hospitals in Benin and Burkina Faso and their surroundings. There were differences in the HWW collection systems in Benin and Burkina Faso in comparison to Finland, which, among other factors, seem to be reflected to some extent in the taxonomical compositions and, therefore, resistomes found in these HWW. However, taxonomy explained the variance in the resistomes from the different countries only partially at least for genus level. Although there were fewer dissimilarities in the resistomes between HWW from Benin and Burkina Faso than in the comparisons with Finland, it is important to consider the differences among West African countries in AMR surveillance.

## 4. Material and Methods

### 4.1. Sample description

Hospital wastewater (HWW) samples were collected in Benin from four hospitals (hospitals A-D, n = 26) and in Burkina Faso from five different hospitals (hospitals E-I, n = 34) in November and December 2019. In Finland, HWW samples were collected from four different hospitals (hospitals J-O, n = 8) in January 2020. The various wards, clinics, and other units typically had their own septic tanks or sumps in the Beninese and Burkinabe hospitals. In Burkina Faso, the samples were mainly from septic tanks or sewers in the hospital area. In Benin, none of the hospitals was connected to a sewer system, and the samples were from septic tanks or sumps (unstructured wastewater wells), which were never emptied to our knowledge. In most cases, the toilet water was not directed into these sumps. For comparison, 11 non-HWW water sources were sampled. These included samples from the tap (n = 1; BENN tap water (drinking)) and river waters (n = 3; BENN river P, Q, R) used for drinking in a remote countryside village in the community of Savalou in central Benin as well as drinking water from a well located in 100 m distance from hospital A in Benin (n = 1; BENN well water A). Street gutter water near hospital B (in a 100 m distance) (n = 1; BENN street gutter A) and a water puddle at hospital C yard next to HWW septic tank (n = 1; BENN puddle at yard C) and a tank distributing water for hand-washing in hospital C in Benin (n = 1; BENN hand-washing C) were also sampled. In Burkina Faso, samples from biologically treated HWW from hospital H (n = 1; BF exit after biological treatment) and a wetland receiving this water (n = 1; BF wetland receiving treated HWW) were collected. Additionally, one sample was collected from wastewater treated in a local WWTP and destined for a river in Burkina Faso (n = 1; BF receiving river after WWTP) (Table 1). Detailed sample descriptions are provided in the Supporting Information, Table S1, and S2. Illustrative pictures and a map indicating sample collection regions are shown in Figures S1 and S2.

### 4.2. DNA extraction and metagenomic sequencing

Water samples were collected into 1-liter bottles and transported to a laboratory on ice. They were kept at +4 °C until processed within 24 hours. A volume of 50–100 ml was filtered through a 0.2 μm polycarbonate filter (Whatman™, GE Healthcare Life Sciences) using a portable vacuum pump (Millivac-Mini Vacuum Pump XF54, Millipore, Merck). DNA was extracted from the filters using the Qiagen Dneasy PowerWater DNA Kit following manufacturer instructions. The concentration and quality of the extracted DNA were determined with a NanoDrop spectrophotometer. Altogether 93 samples were subjected to shotgun metagenomic sequencing using Illumina Novaseq5000 with Nextera XT library preparation at the Institute of Biotechnology, University of Helsinki.

### 4.3. Bioinformatic analyses

All quality control and read mapping analyses were run using an in-house Snakemake ^70^ (v 5.3.0) workflow. Briefly, the quality control steps included in the workflow were performed using FastQC ^71^ (v0.11.8) and MultiQC ^72^ (v1.9), with adapter and low-quality read removal using Cutadapt ^73^ (v2.7) (parameters -O 10 -m 30 -q 20). Nucleotide sequence reads were mapped using Bowtie2 ^74^ (v 2.4.1) (parameters -D 20 -R 3 -N 1 -L 20 -I S,1,0.50) against the ResFinder database ^75^ (version 3.2, downloaded on 2020-06-28). The reads were sorted and filtered using SAMtools ^76^ (v1.9) such that the reads mapping as pair or alone were calculated as a single count. Mobile genetic elements were identified by mapping the reads with Bowtie2 similarly as above against the MobileGeneticElementDatabase ^58^ (https://github.com/KatariinaParnanen/MobileGeneticElementDatabase, downloaded on 2020-06-28) consisting of 2714 unique MGE sequences, including transposons; integrons of classes 1, 2, and 3; the integron-associated disinfectant resistance gene *qacEτ1*.; and IS and ISCR type insertion sequences.

Taxonomic profiling was performed using both Metaphlan3 ^77^ (v3.0.1) and Metaxa2 ^78^ (v.2.2.1). Metaphlan3 was run to achieve both relative abundances and ‘absolute abundances’ defined by the program developers using the parameter ‘-t rel_ab_w_read_stats’. The outputs from the former were used for ordinations and the latter for diversity analyses. In both cases, the merged abundance tables were modified so that only taxa which were identified to species level were included in the downstream analyses. However, knowing the limitations of species-level taxa identification using shotgun metagenomics ^79^, the results were presented only at the genus level after taxa agglomeration by the function ‘tax_glom’ from the ‘phyloseq’ package ^80^ (v1.40.0) prior analyses.

The counts for bacterial 16S rRNA from Metaxa2 were used to normalize ARG and MGE counts to obtain relative abundances. Gene lengths were taken into account in the normalization. Three HWW samples were collected in duplicate (Table S2). These replicates were not included in the statistical analyses, but they were used to evaluate the selected methods for detecting resistance. The sums of the relative abundances of ARGs in the replicated sample pairs were highly similar to each other (Figure S10).

All resistance genes found in the ResFinder database ^75^ were clustered based on 90 % similarity in their sequence identity using CD-HIT ^39^ (v4.8.1). This clustering was utilized to determine the groups of *bla*OXA-48- and *bla*OXA-58-like variants for studying carbapenemase genes. Data available in Beta-Lactamase DataBase (BLDB) ^41^ was used to confirm the carbapenemase activity of the variants of *bla*OXA-48 and *bla*OXA-58-like clusters as well as other carbapenemases studied here (Table S6). For instance, only *bla*GES variants known to encode carbapenemases were included in the visualization (Figure 3), while those encoding ESBL phenotypes were not. Similarly, ARG clustering of 90 % shared nucleotide identity was applied to group *mcr* variants (Table S7).

To study the genetic environment of ARGs, a subset of samples was assembled into contigs with MEGAHIT ^81^ (v1.2.8) with parameters --min-contig-len 1000 -m 32000000000. Anvi’o ^82^ (v7) and Bandage ^83^ programs were applied to visualize the genetic environments (Figures S5 and S7, see details in Supporting Information 1). For *mcr-5* gene, one to two *mcr-5* positive samples from all studied countries were selected for the assembly and the analysis of the genetic background. The same samples were used for the search of putative integron carried multi-drug resistance gene cassettes. Those putative gene cassettes that had multiple ARGs encoded by the same fragment were represented in the Figure S5.

### 4.4. Statistical analyses

All statistical analyses described below were performed for the 68 HWW samples (Table 1, Table S1). The sum of the relative abundances for ARGs, MGEs, and *intI1* was obtained using 16S rRNA gene counts and gene lengths to normalize the count data. As the data did not fulfill the assumptions of normality, a Kruskal-Wallis test from the ‘stats’ package ^84^ (v 4.2.0) was applied to study whether the differences between countries were significant. Pair-wise, Wilcoxon rank sum tests adjusted by Benjamini-Hochberg from the ‘stats’ package ^84^ (v 4.2.0) were performed to determine which comparisons were significant for ARGs and *intI1*.

#### 4.4.1. Compositional analyses

Next-generation sequencing (NGS) data is compositional as it only contains relative information ^43^. By ignoring this compositional nature of NGS data, results conducted by traditional normalization methods may suffer from technical artifacts due to sequencing depth limitations ^85^. ANOVA-Like Differential Expression tool for high throughput sequencing data (ALDEx2) ^38^ (v1.28.0) was applied to study the divergent features of the resistomes in HWW from each country Benin, Burkina Faso, and Finland. ALDEx2 handles the compositionality of the data by applying suitable data transformations and expressing the results of feature abundance values relative to the geometric mean abundance of other features in a sample or a selected reference gene. Here, for ARGs, the 16S rRNA gene was used as the reference for additive alr-transformation. First, the ARG count data was split so that pair-wise comparisons between countries were possible (e.g., Benin vs. Finland). The command ‘aldex.clr(df, conditions, denom = ref)’ excludes the features with zero count in all samples and performs the alr-transformation with the selected reference gene. The significance of the comparisons was tested using the Wilcoxon rank test (command ‘aldex.ttest(x, paired.test = FALSE, verbose = FALSE)’), in which Benjamini-Hochberg corrections control false positive identifications. Finally, the effect sizes and the within and between condition values were estimated with the command ‘aldex.effect’. ALDEx2 ^38^ was also applied to study the differentially abundant taxa between the HWW from different countries. Metaphlan3 count data (generated using the parameter ‘-t rel_ab_w_read_stats’) was clr-transformed in the analysis, and the significance of the comparisons was tested similarly as above.

For PCA ordinations, clr-transformed ARG, MGE and taxa counts (generated by Metaphlan3 with parameter ‘-t rel_ab_w_read_stats’) were visualized using ‘microViz’ ^86^ (v0.9.1) with the command ‘count_data %>% tax_transform(“clr”) %>% ord_calc(method = “PCA”) %>% ord_plot(color = “country”‘. For PCA taxa were fixed to genus level as described earlier.

All statistical analyses were performed in RStudio (v4.2.0), and the results were visualized using ‘ggplot2’ ^87^ (v3.3.6), and ‘patchwork’ ^88^ (v1.1.1). Vector maps were drawn using the package ‘rnaturalearth’ ^89^ (v0.1.0), ‘ggspatial’ ^90^ (v1.1.6), and ‘maps’ ^91^ (v3.4.0).

## Data Availability

The data for this study have been deposited in the European Nucleotide Archive (ENA) at EMBL-EBI under the accession number PRJEB47975 and will be public upon article publication in a peer reviewed journal.

## Ethical permission

The ethical permissions for the project were received from ‘Comité National d’Ethique pour la Recherche en Santé’ under the Health Ministry in Benin and ‘Comité d’Ethique pour la Recherche en Santé’ under the Health Ministry in Burkina Faso.

## Supporting Information

The supporting information referred to within the text as Figures S1-S11 and Tables S1-S10 can be found in the ‘Supporting Information 1’ (Figures S1-S11) and ‘Supporting Information 2’ (Tables S1-S10).

## Acknowledgments

We thank the staff in all the participating hospitals for their collaboration in sampling. We also thank the laboratory personnel of the URMAPha at the University of Abomey-Calavi in Benin and the Clinical Research Unit of Nanoro (CRUN) in Burkina Faso for sample processing. Finally, we acknowledge CSC, IT Center for Science, Finland, for providing the computational resources for the study.

## Data availability

The data for this study have been deposited in the European Nucleotide Archive (ENA) at EMBL-EBI under the accession number PRJEB47975 and will be public upon article publication in a peer-reviewed journal.

## Code availability

All custom codes used for the analyses are available from https://github.com/melinamarkkanen/AMRIWA upon article publication in a peer-reviewed journal. Analysis scripts are provided for the reviewers prior publication.

## References

1. World Health Organization. Global Action Plan on Antimicrobial Resistance. Published online 2015. doi:10.13140/RG.2.2.24460.77448

2. Ventola CL. The Antibiotic Resistance Crisis Part 1: Causes and Threats. Physichal Therapy. 2015;40(4):277–283.

3. Laxminarayan R, Duse A, Wattal C, et al. Antibiotic resistance-the need for global solutions. Lancet Infectious Diseases. 2013;13(12):1057–1098. doi:10.1016/S1473-3099(13)70318-9

4. Jacobs J, Hardy L, Semret M, et al. Diagnostic Bacteriology in District Hospitals in Sub-Saharan Africa: At the Forefront of the Containment of Antimicrobial Resistance. Front Med (Lausanne). 2019;6:205. doi:10.3389/fmed.2019.00205

5. Belachew SA, Hall L, Selvey LA. Non-prescription dispensing of antibiotic agents among community drug retail outlets in Sub-Saharan African countries: a systematic review and meta-analysis. Antimicrob Resist Infect Control. 2021;10(1). doi:10.1186/s13756-020-00880-w

6. Butcher A, Cañada JA, Sariola S. How to make noncoherent problems more productive: Towards an AMR management plan for low resource livestock sectors. Humanit Soc Sci Commun. 2021;8(1):287. doi:10.1057/s41599-021-00965-w

7. Van TTH, Yidana Z, Smooker PM, Coloe PJ. Antibiotic use in food animals worldwide, with a focus on Africa: Pluses and minuses. J Glob Antimicrob Resist. 2020;20:170–177. doi:10.1016/j.jgar.2019.07.031

8. Martínez JL, Coque TM, Baquero F. What is a resistance gene? Ranking risk in resistomes. Nat Rev Microbiol. 2014;13:116–123. doi:10.1038/nrmicro3399

9. Zhang AN, Gaston JM, Dai CL, et al. An omics-based framework for assessing the health risk of antimicrobial resistance genes. Nat Commun. 2021;4765. doi:10.1038/s41467-021-25096-3

10. Nielsen TK, Browne PD, Hansen LH. Antibiotic resistance genes are differentially mobilized according to resistance mechanism. Gigascience. 2022;11. doi:10.1093/gigascience/giac072

11. Karkman A, Berglund F, Flach CF, Kristiansson E, Larsson DGJ. Predicting clinical resistance prevalence using sewage metagenomic data. Commun Biol. 2020;3(1):711. doi:10.1038/s42003-020-01439-6

12. Gillings MR, Gaze WH, Pruden A, Smalla K, Tiedje JM, Zhu YG. Using the class 1 integron-integrase gene as a proxy for anthropogenic pollution. ISME J. 2015;9:1269–1279. doi:10.1038/ismej.2014.226

13. Li Y, Yang X, Zhang J, et al. Molecular characterisation of antimicrobial resistance determinants and class 1 integrons of Salmonella enterica subsp. enterica serotype Enteritidis strains from retail food in China. Food Control. 2021;128:108191. doi:10.1016/j.foodcont.2021.108191

14. Gillings MR. Integrons: Past, Present, and Future. Microbiology and Molecular Biology Reviews. 2014;78(2):257–277. doi:10.1128/MMBR.00056-13

15. World Health Organization. Global Antimicrobial Resistance and Use Surveillance System (GLASS) Report 2021. Geneva.; 2021.

16. World Health Organization. WHO report on surveillance of antibiotic consumption: 2016-2018 early implementation. Published 2018. https://www.who.int/medicines/areas/rational_use/oms-amr-amc-report-2016-2018/en/

17. Suay-García, Pérez-Gracia. Present and Future of Carbapenem-resistant Enterobacteriaceae (CRE) Infections. Antibiotics. 2019;8(3):122. doi:10.3390/antibiotics8030122

18. Poirel L, Marqué S, Héritier C, Segonds C, Chabanon G, Nordmann P. OXA-58, a novel class D β-lactamase involved in resistance to carbapenems in Acinetobacter baumannii. Antimicrob Agents Chemother. 2005;49(1):202–208. doi:10.1128/AAC.49.1.202-208.2005

19. Poirel L, Weldhagen GF, Naas T, de Champs C, Dove MG, Nordmann P. GES-2, a class A β-lactamase from Pseudomonas aeruginosa with increased hydrolysis of imipenem. Antimicrob Agents Chemother. 2001;45(9):2598–2603. doi:10.1128/AAC.45.9.2598-2603.2001

20. Nordmann P, Poirel L. The difficult-to-control spread of carbapenemase producers among Enterobacteriaceae worldwide. Clinical Microbiology and Infection. 2013;20:821–830. doi:10.1111/1469-0691.12719

21. Falagas ME, Kasiakou Sofia. K. Colistin: The Revival of Polymyxins for the Management of Multidrug-Resistant Gram-Negative Bacterial Infections. Clinical Infectious Diseases. 2005;40(9):1333–1341.

22. Ling Z, Yin W, Shen Z, Wang Y, Shen J, Walsh TR. Epidemiology of mobile colistin resistance genes mcr-1 to mcr-9. Journal of Antimicrobial Chemotherapy. 2020;75(11):3087–3095. doi:10.1093/jac/dkaa205

23. Anyanwu MU, Odilichukwu C, Okpala R, Chah KF, Shoyinka VS. Prevalence and Traits of Mobile Colistin Resistance Gene Harbouring Isolates from Different Ecosystems in Africa. Biomed Res Int. 2021;22:6630379. doi:10.1155/2021/6630379

24. Collignon P, Beggs JJ, Walsh TR, Gandra S, Laxminarayan R. Anthropological and socioeconomic factors contributing to global antimicrobial resistance: a univariate and multivariable analysis. Lancet Planet Health. 2018;2(9):e398–405.

25. van Boeckel TP, Pires J, Silvester R, et al. Global trends in antimicrobial resistance in animals in low- and middle-income countries. Science (1979). 2019;365(6459):eaaw1944. doi:10.1126/science.aaw1944

26. Iskandar K, Molinier L, Hallit S, et al. Surveillance of antimicrobial resistance in low- and middle-income countries: a scattered picture. Antimicrob Resist Infect Control. 2020;10:63. doi:10.1186/s13756-021-00931-w

27. Ambler RP. The structure of beta-lactamases. Philos Trans R Soc Lond. 1980;289(1036):321–331. doi:10.1098/rstb.1980.0049

28. Bernabé KJ, Langendorf C, Ford N, Ronat JB, Murphy RA. Antimicrobial resistance in West Africa: a systematic review and meta-analysis. Int J Antimicrob Agents. 2017;50(5):629–639. doi:10.1016/j.ijantimicag.2017.07.002

29. Mahamat OO, Kempf M, Lounnas M, et al. Epidemiology and prevalence of extended-spectrum β-lactamase-and carbapenemase-producing Enterobacteriaceae in humans, animals and the environment in West and Central Africa. Int J Antimicrob Agents. 2021;57:106203. doi:10.1016/j.ijantimicag.2020.106203

30. Hendriksen RS, Munk P, Njage P, et al. Global monitoring of antimicrobial resistance based on metagenomics analyses of urban sewage. Nat Commun. 2019;10(1):1124. doi:10.1038/s41467-019-08853-3

31. Pärnänen KMM, Narciso-da-Rocha C, Kneis D, et al. Antibiotic resistance in European wastewater treatment plants mirrors the pattern of clinical antibiotic resistance prevalence. Sci Adv. 2019;5(3):eaau9124. doi:10.1126/sciadv.aau9124

32. European Centre for Disease Prevention and Control. Antimicrobial Resistance in the EU/EEA (EARS-Net) - Annual Epidemiological Report 2019.; 2020. https://www.ecdc.europa.eu/en/publications-data/surveillance-antimicrobial-resistance-europe-2019

33. Buelow E, Rico A, Gaschet M, et al. Hospital discharges in urban sanitation systems: Long-term monitoring of wastewater resistome and microbiota in relationship to their eco-exposome. Water Res X. 2020;7:100045. doi:10.1016/j.wroa.2020.100045

34. Karkman A, Do TT, Walsh F, Virta MPJ. Antibiotic-Resistance Genes in Waste Water. Trends Microbiol. 2018;26(3):220–228. doi:10.1016/J.TIM.2017.09.005

35. Luk-in S, Pulsrikarn C, Bangtrakulnonth A, Chatsuwan T, Kulwichit W. Occurrence of a novel class 1 integron harboring qnrVC4 in Salmonella Rissen. Diagn Microbiol Infect Dis. 2017;88(3):282–286. doi:10.1016/j.diagmicrobio.2017.03.016

36. Xia R, Guo X, Zhang Y, Xu H. qnrVC-like gene located in a novel complex class 1 integron harboring the ISCR1 element in an Aeromonas punctata strain from an aquatic environment in Shandong Province, China. Antimicrob Agents Chemother. 2010;54(8):3471–3474. doi:10.1128/AAC.01668-09

37. Fernandes AD, Reid JNS, Macklaim JM, McMurrough TA, Edgell DR, Gloor GB. Unifying the analysis of high-throughput sequencing datasets: Characterizing RNA-seq, 16S rRNA gene sequencing and selective growth experiments by compositional data analysis. Microbiome. 2014;2(1). doi:10.1186/2049-2618-2-15

38. Fernandes AD, Reid JN, Macklaim JM, Mcmurrough TA, Edgell DR, Gloor GB. Unifying the analysis of high-throughput sequencing datasets: characterizing RNA-seq, 16S rRNA gene sequencing and selective growth experiments by compositional data analysis. Microbiome. 2014;2:15.

39. Fu L, Niu B, Zhu Z, Wu S, Li W. Sequence analysis CD-HIT: accelerated for clustering the next-generation sequencing data. Bioinformatics. 2012;28(23):3150–3152. doi:10.1093/bioinformatics/bts565

40. Ma L, Li AD, Yin X le, Zhang T. The Prevalence of Integrons as the Carrier of Antibiotic Resistance Genes in Natural and Man-Made Environments. Environ Sci Technol. 2017;51(10):5721–5728. doi:10.1021/acs.est.6b05887

41. Naas T, Oueslati S, Bonnin RA, et al. Beta-lactamase database (BLDB)-structure and function. J Enzyme Inhib Med Chem. 2017;32(1):917–919. doi:10.1080/14756366.2017.1344235

42. Rice EW, Wang P, Smith AL, Stadler LB. Determining Hosts of Antibiotic Resistance Genes: A Review of Methodological Advances. Environ Sci Technol Lett. 2020;7(5):282–291. doi:10.1021/acs.estlett.0c00202

43. Aitchison J. The Statistical Analysis of Compositional Data. Journal of the Royal Statistical Society: Series B (Methodological). 1982;44(2):139–160.

44. Love MI, Huber W, Anders S. Moderated estimation of fold change and dispersion for RNA-seq data with DESeq2. Genome Biol. 2014;15:550. doi:10.1186/s13059-014-0550-8

45. Araújo S, Sousa M, Tacão M, et al. Carbapenem-resistant bacteria over a wastewater treatment process: Carbapenem-resistant Enterobacteriaceae in untreated wastewater and intrinsically-resistant bacteria in final effluent. Science of the Total Environment. 2021;782:146892. doi:10.1016/j.scitotenv.2021.146892

46. Zielinski W, Korzeniewska E, Harnisz M, Drzymala J, Felis E, Bajkacz S. Wastewater treatment plants as a reservoir of integrase and antibiotic resistance genes – An epidemiological threat to workers and environment. Environ Int. 2021;156:106641. doi:10.1016/j.envint.2021.106641

47. Figueiredo S, Bonnin RA, Poirel L, Duranteau J, Nordmann P. Identification of the naturally occurring genes encoding carbapenem-hydrolysing oxacillinases from Acinetobacter haemolyticus, Acinetobacter johnsonii, and Acinetobacter calcoaceticus. Clinical Microbiology and Infection. 2012;18:907–913. doi:10.1111/j.1469-0691.2011.03708.x

48. Périchon B, Goussard S, Walewski V, et al. Identification of 50 class D β-lactamases and 65 Acinetobacter-derived cephalosporinases in Acinetobacter spp. Antimicrob Agents Chemother. 2014;58(2):936–949. doi:10.1128/AAC.01261-13

49. Olalekan A, Onwugamba F, Iwalokun B, Mellmann A, Becker K, Schaumburg F. High proportion of carbapenemase-producing Escherichia coli and Klebsiella pneumoniae among extended-spectrum β-lactamase-producers in Nigerian hospitals. J Glob Antimicrob Resist. 2020;21:8–12. doi:10.1016/j.jgar.2019.09.007

50. Manenzhe RI, Zar HJ, Nicol MP, Kaba M. The spread of carbapenemase-producing bacteria in Africa: a systematic review. Journal of Antimicrobial Chemotherapy. 2015;70:23–40. doi:10.1093/jac/dku356

51. van Duin D, Doi Y. The global epidemiology of carbapenemase-producing Enterobacteriaceae. Virulence. 2017;8(4):460–469. doi:10.1080/21505594.2016.1222343

52. Mushi MF, Mshana SE, Imirzalioglu C, Bwanga F. Carbapenemase Genes among Multidrug Resistant Gram Negative Clinical Isolates from a Tertiary Hospital in Mwanza, Tanzania. Biomed Res Int. 2014;2014(4):303104. doi:10.1155/2014/303104

53. WHO Regional Office for Europe and European Centre for Disease Prevention and Control. Surveillance of Antimicrobial Resistance in Europe, 2020. Executive Summary. Copenhagen: WHO Regional for Europe.; 2021.

54. Räisänen K, Lyytikäinen O, Kauranen J, et al. Molecular epidemiology of carbapenemase-producing Enterobacterales in Finland, 2012–2018. European Journal of Clinical Microbiology and Infectious Diseases. 2020;39(9):1651–1656. doi:10.1007/S10096-020-03885-W

55. Yatsunenko T, Rey FE, Manary MJ, et al. Human gut microbiome viewed across age and geography. Nature. 2012;486:222–227. doi:10.1038/nature11053

56. Thompson L, Sanders JG, McDonald D, et al. A communal catalogue reveals Earth’s multiscale microbial diversity. Nature. 2017;551(18):457–463. doi:10.1038/nature24621

57. Oyem IM, Oyem HH, Atuanya. Molecular characterisation of bacteria strains of septic tank sewage samples from related sites in Delta and Edo States of Nigeria using 16S rRNA denaturing gradient gel electrophoresis (DGGE). Afr J Environ Sci Tech. 2020;14(6):132–138. doi:10.5897/AJEST2020.2851

58. Pärnänen K, Karkman A, Hultman J, et al. Maternal gut and breast milk microbiota affect infant gut antibiotic resistome and mobile genetic elements. Nat Commun. 2018;9:389. doi:10.1038/s41467-018-06393-w

59. Olumuyiwa Olaitan A, Dandachi I, Alexandra Baron S, Daoud Z, Morand S, Rolain JM. Banning colistin in feed additives: a small step in the right direction. Lancet Infect Dis. 2021;21(1):29–30. doi:10.1016/S1473

60. Olowo-Okere A, Yacouba A. Review Molecular mechanisms of colistin resistance in Africa: A systematic review of literature. Germs. 2020;10(4):367–379.

61. Kneis D, Berendonk TU, Heß S. High prevalence of colistin resistance genes in German municipal wastewater. Science of the Total Environment. 2019;694:133454. doi:10.1016/j.scitotenv.2019.07.260

62. Ngbede EO, Poudel A, Kalalah A, et al. Identification of mobile colistin resistance genes (mcr-1.1, mcr-5 and mcr-8.1) in Enterobacteriaceae and Alcaligenes faecalis of human and animal origin, Nigeria. Int J Antimicrob Agents. 2020;56(3):106108. doi:10.1016/j.ijantimicag.2020.106108

63. Borowiak M, Fischer J, Hammerl JA, Hendriksen RS, Szabo I, Malorny B. Identification of a novel transposon-associated phosphoethanolamine transferase gene, mcr-5, conferring colistin resistance in d-tartrate fermenting Salmonella enterica subsp. enterica serovar Paratyphi B. Journal of Antimicrobial Chemotherapy. 2017;72(12):3317–3324. doi:10.1093/jac/dkx327

64. Borowiak M, Hammerl JA, Deneke C, Fischer J, Szabo I, Malorny B. Characterization of mcr-5-Harboring Salmonella enterica subsp. enterica Serovar Typhimurium Isolates from Animal and Food Origin in Germany. Antimicrob Agents Chemother. 2019;63(6):e00063–19. doi:10.1128/AAC.00063-19

65. Guo S, Tay MYF, Thu AK, et al. Conjugative IncX1 plasmid harboring colistin resistance gene mcr-5.1 in Escherichia coli isolated from chicken rice retailed in Singapore. Antimicrob Agents Chemother. 2019;63(11):e01043–19. doi:10.1128/AAC.01043-19

66. Peace Hounkpe S, Codjo Adjovi E, Crapper M, Awuah E. Wastewater Management in Third World Cities: Case Study of Cotonou. Benin Journal of Environmental Protection. 2014;5(05):387–399. doi:10.4236/jep.2014.55042

67. Totin HS, Ernest A, Odoulami L, Edorh PA, Boukari M, Boko M. Groundwater pollution and the safe water supply challenge in Cotonou town, Benin (West Africa). IAHS-AISH Proceedings and Reports. 2013;361:191–196. doi:10.13140/RG.2.1.2668.0168

68. Bougnom BP, Zongo C, McNally A, et al. Wastewater used for urban agriculture in West Africa as a reservoir for antibacterial resistance dissemination. Environ Res. 2019;168:14–24. doi:10.1016/j.envres.2018.09.022

69. Bougnom BP, Thiele-Bruhn S, Ricci V, Zongo C, Piddock LJV. Raw wastewater irrigation for urban agriculture in three African cities increases the abundance of transferable antibiotic resistance genes in soil, including those encoding extended spectrum β-lactamases (ESBLs). Science of the Total Environment. 2020;698:134201. doi:10.1016/j.scitotenv.2019.134201

70. Mölder F, Jablonski KP, Letcher B, et al. Sustainable data analysis with Snakemake. F1000Res. 2021;10:33. doi:10.12688/f1000research.29032.1

71. Andrews S. FastQC A quality control tool for high throughput sequence data https://www.bioinformatics.babraham.ac.uk/projects/fastqc/.

72. Ewels P, Ns Magnusson M, Lundin S, Aller MK. Data and text mining MultiQC: summarize analysis results for multiple tools and samples in a single report. Bioinformatics. 32(19):3047–3048. doi:10.1093/bioinformatics/btw354

73. Marcel M. Cutadapt removes adapter sequences from high-throughput sequencing reads. EMBnet J. 2011;17(1):10–12. doi:http://dx.doi.org/10.14806/ej.17.1.200

74. Langmead B, Salzberg SL. Fast gapped-read alignment with Bowtie 2. Nat Methods. 2012;9(4):357–359. doi:10.1038/nmeth.1923

75. Bortolaia V, Kaas RS, Ruppe E, et al. ResFinder 4.0 for predictions of phenotypes from genotypes. Journal of Antimicrobial Chemotherapy. 2020;75(12):3491–3500. doi:10.1093/jac/dkaa345

76. Li H, Handsaker B, Wysoker A, et al. The Sequence Alignment/Map format and SAMtools. Bioinformatics. 2009;25(16):2078–2079. doi:10.1093/bioinformatics/btp352

77. Beghini F, Mciver LJ, Blanco-mi A, et al. Integrating taxonomic, functional, and strain-level profiling of diverse microbial communities with bioBakery 3. Elife. 2021;4(10):1–42.

78. Bengtsson-Palme J, Hartmann M, Eriksson KM, et al. metaxa2: Improved identification and taxonomic classification of small and large subunit rRNA in metagenomic data. Mol Ecol Resour. 2015;15(6):1403–1414. doi:10.1111/1755-0998.12399

79. Thomas AM, Segata N. Multiple levels of the unknown in microbiome research. BMC Biol. 2019;17(48). doi:10.1186/s12915-019-0667-z

80. McMurdie PJ, Holmes S. Phyloseq: An R Package for Reproducible Interactive Analysis and Graphics of Microbiome Census Data. PLoS One. 2013;8(4). doi:10.1371/journal.pone.0061217

81. Li D, Liu CM, Luo R, Sadakane K, Lam TW. MEGAHIT: an ultra-fast single-node solution for large and complex metagenomics assembly via succinct de Bruijn graph. Bioinformatics. 2015;31(10):1674–1676. doi:10.1093/bioinformatics/btv033

82. Murat Eren A, Kiefl E, Shaiber A, et al. Community-led, integrated, reproducible multi-omics with anvi’o. Nat Microbiol. 2021;6:3–6. doi:10.1038/s41564-020-00834-3

83. Wick RR, Schultz MB, Zobel J, Holt KE. Bandage: interactive visualization of de novo genome assemblies. Bioinformatics. 2015;31(20):3350–3352. doi:10.1093/bioinformatics/btv383

84. R Foundation for Statistical Computing. Vienna. Austria. R: A language and environment for statistical computing. R Foundation for Statistical Computing, Vienna, Austria. :https://www.R-project.org/. https://www.r-project.org/.

85. Quinn TP, Erb I, Gloor G, Notredame C, Richardson MF, Crowley TM. A field guide for the compositional analysis of any-omics data. Gigascience. 2019;8(9). doi:10.1093/gigascience/giz107

86. Barnett D, Arts I, Penders J. microViz: an R package for microbiome data visualization and statistics. J Open Source Softw. 2021;6(63):3201. doi:10.21105/joss.03201

87. Wickham H. ggplot2: Elegant graphics for data analysis. https://ggplot2.tidyverse.org.

88. Pedersen TL. patchwork: The Composer of Plots. https://cran.r-project.org/package=patchwork.

89. South A. rnaturalearth: World Map Data from Natural Earth. https://cran.r-project.org/package=rnaturalearth.

90. Dunnington D. ggspatial: Spatial Data Framework for ggplot2. https://paleolimbot.github.io/ggspatial/.

91. Becker, Minka Deckmyn. maps: Draw Geographical Maps. https://cran.r-project.org/package=maps.

