## Supporting Information 1: Figures S1-S11 for "Metagenomic analysis of the abundance and composition of antibiotic resistance genes in hospital wastewater in Benin, Burkina Faso, and Finland"

Table of Contents:

1. Sample Collection
   **Figure S1.** Sample collection in Benin and Burkina Faso (p. 2)
   **Figure S2.** Examples of sampling sites (p. 3)
2. Supporting Results
   **Figure S3.** Diversity of ARGs, MGEs, and taxa (p. 4)
   **Figure S4.** Correlation between ARGs and MGEs (p. 6)
   **Figure S5.** Examples of putative class 1 integron elements with ARGs (p. 7)
   **Figure S6.** PCA plots for ARGs, MGEs, and taxa with all collected samples (p. 8)
   **Figure S7.** Examples of genetic environments of *mcr-5* genes (p. 10)
   **Figure S8.** Significantly differentially abundant taxa in HWW from Benin, Burkina Faso, and Finland (p. 11)
   **Figure S9.** Top 11 most abundant taxa in all samples (p. 12)
   **Figure S10.** Analysis of replicate samples (p. 13)
   **Figure S11.** Correlations between ALDEx2 and DESeq2 results for significantly differentially abundant ARGs (p. 14)
3. References (p. 14)

Pages: 15
Figures: 11

1. Sample collection

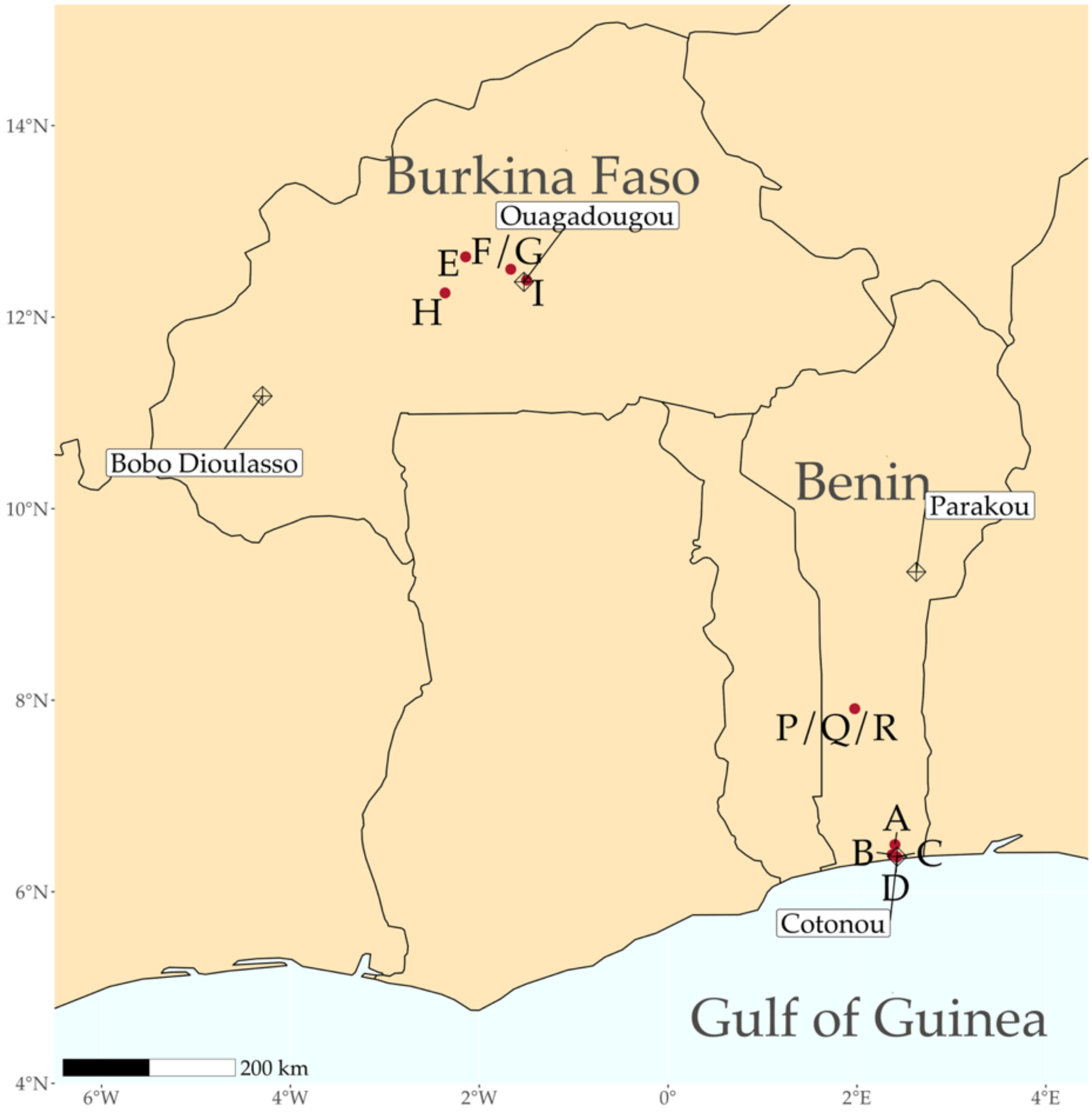


**Figure S1.** Sample collection in Benin and Burkina Faso. Sample collection sites are denoted with red dots on the map. The climate in the coastal region of Benin is hot and humid. The average daytime temperature in Cotonou varies between 28–32 °C and humidity between 77–84 %, with an annual rainfall of 1,308 mm. Burkina Faso is characterized by a dry tropical climate. The average daytime temperature in Ouagadougou varies between 31–39 °C, and the annual rainfall is 745 mm. During the sampling period in November/December, it typically rains only a little in Cotonou and not at all in Ouagadougou.

A) 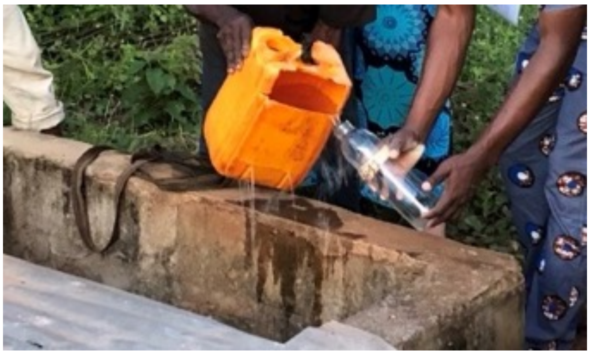 B) 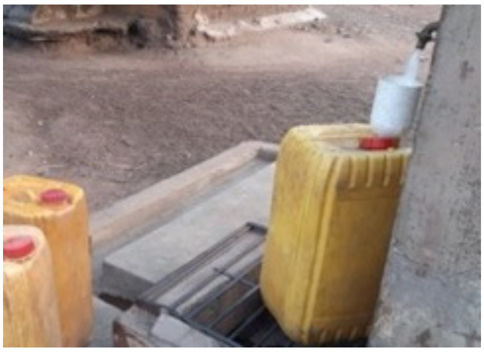

C) 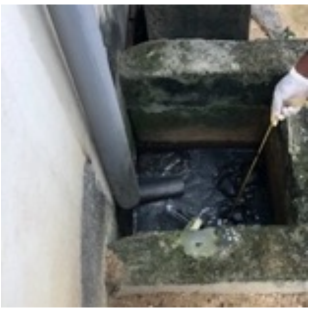 D) 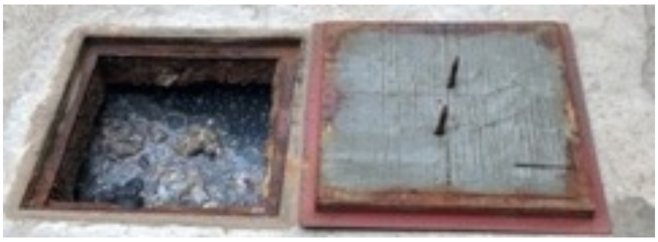

E) 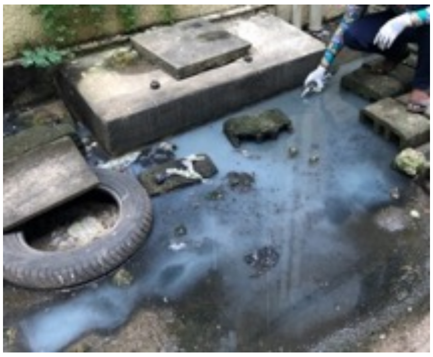 F) 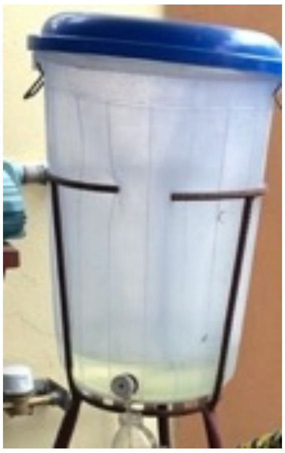 G) 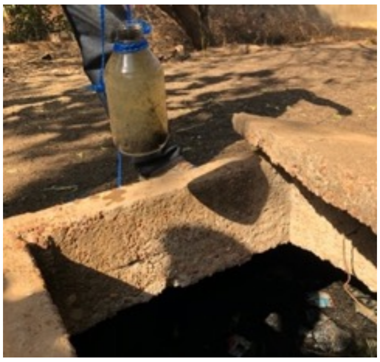

**Figure S2.** Examples of sampling sites. The samples collected in Benin included A) well water 100 m away from hospital A in Benin (sample BH11; BENN well water A); B) tap water in a distant village in Benin (sample BSE93; BENN tap water (drinking)); C) HWW from an open cesspit next to the wall at the hospital B (sample BH29; BENN HWW B); D) the central septic tank collecting wastewater from different sections of hospital C (sample BH44; BENN HWW C); E) water puddle surrounding the surgery room septic tank at hospital C (sample BH48; BENN puddle at yard C); F) water from a tank used for washing hands at hospital C (sample BH52; BENN hand-washing C). G) In Burkina Faso, the HWW septic tank is shown for hospital E (sample BFH1; BF HWW E). Photos by K. Haukka, A. Sarekoski and A. Butcher.

### 2. Supporting results

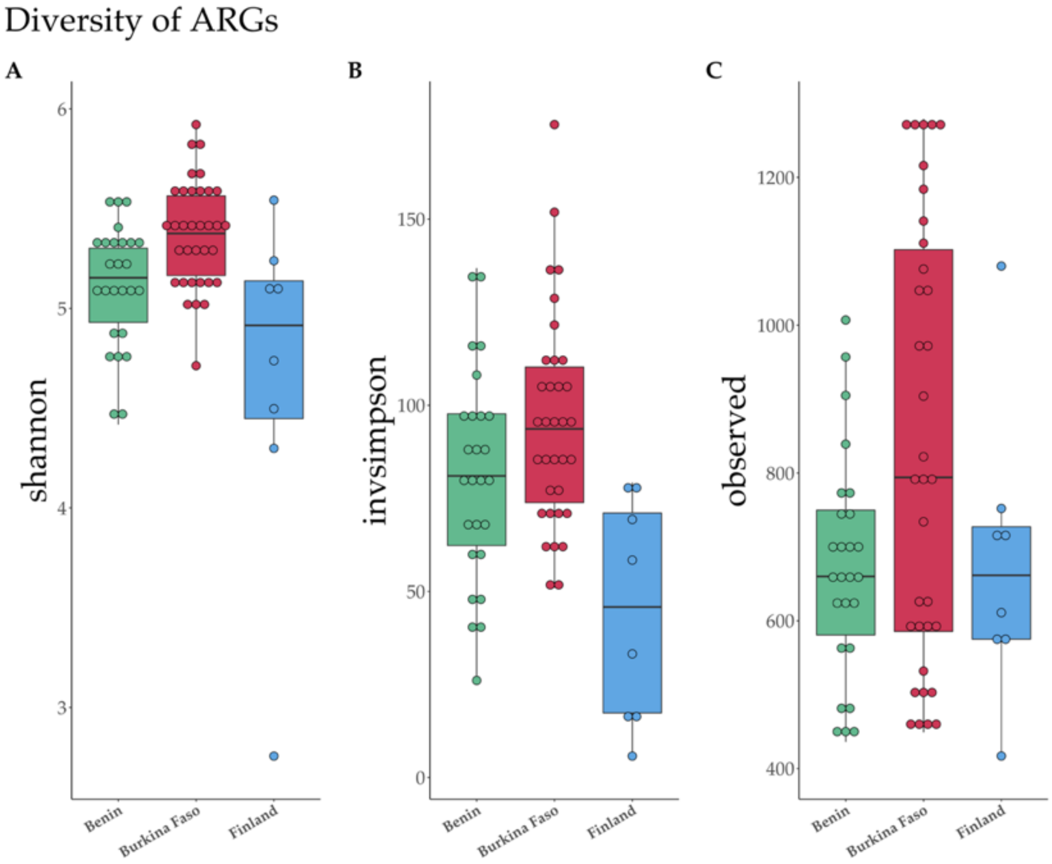
Diversity of ARGs
A
B
C
shannon
invsimpson
observed
6
5
4
3
150
100
50
0
1200
1000
800
600
400
Benin
Burkina Faso
Finland

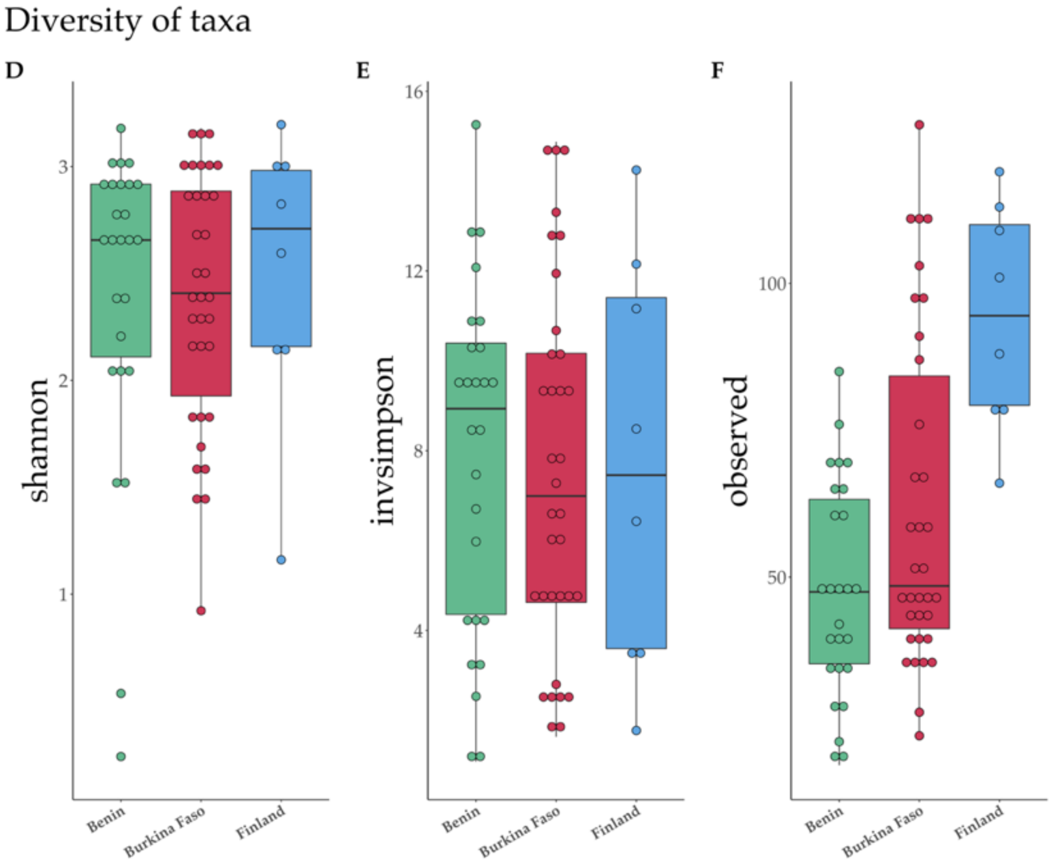
Diversity of taxa
D
E
F
shannon
invsimpson
observed
3
2
1
16
12
8
4
100
50
Benin
Burkina Faso
Finland

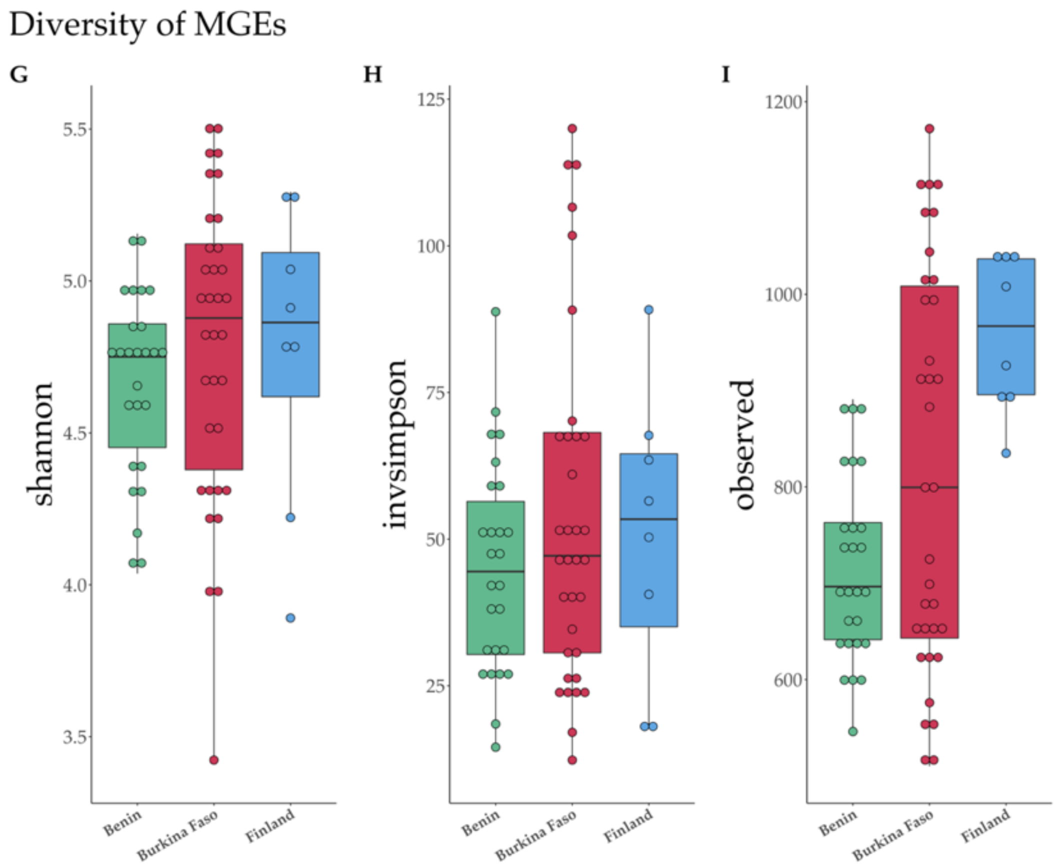


**Figure S3.** Diversity indexes (Shannon, Inverse Simpson, and Observed) for ARGs (A-C), taxa (D-F), and MGEs (G-I). In the boxplots, country medians are shown as vertical lines and the interquartile ranges (25th and 75th percentiles) as boxplot hinges. The horizontal lines represent the highest and lowest values. Outliers are values higher or below 1.5 times the upper or lower percentiles, respectively. The outlier samples were FH6 (A), BFH35 (B), FH1 (C), BH01 and BH09 (D), and FH6 (G). Based on pairwise Wilcoxon rank sum tests adjusted by Benjamini-Hochberg, the significant ($p < 0.05$) comparisons were: A) Finland – Burkina Faso and Benin – Burkina Faso; B) Finland – Benin and Finland – Burkina Faso; F) Finland – Benin and Finland – Burkina Faso; I) Benin – Finland. Diversity analyses of ARGs, taxa, and MGE were done with the function ‘diversity’ from the ‘vegan’ package [1] (v 2.6.2). Pairwise Wilcoxon rank sum tests adjusted by Benjamini-Hochberg from the ‘stats’ package (78) (v 4.2.0) were performed to determine the significant comparisons.

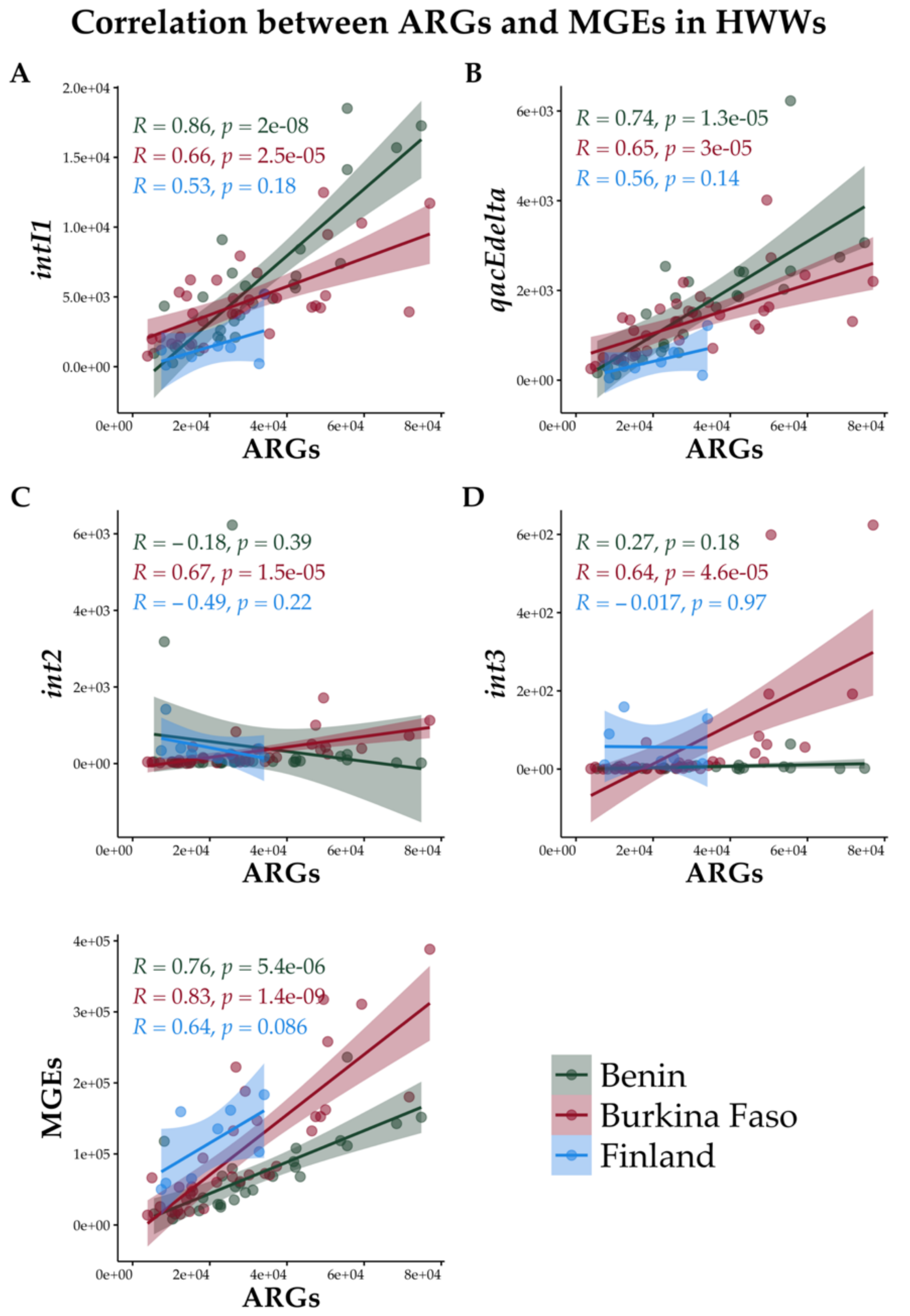


**Figure S4.** Pearson correlations between ARGs and A) *intI1*, B) *qacEΔ*, C) *int2*, and D) *int3* genes, and E) all MGEs. Correlations were computed using package ggpubr [2] (v 0.4.0.999).

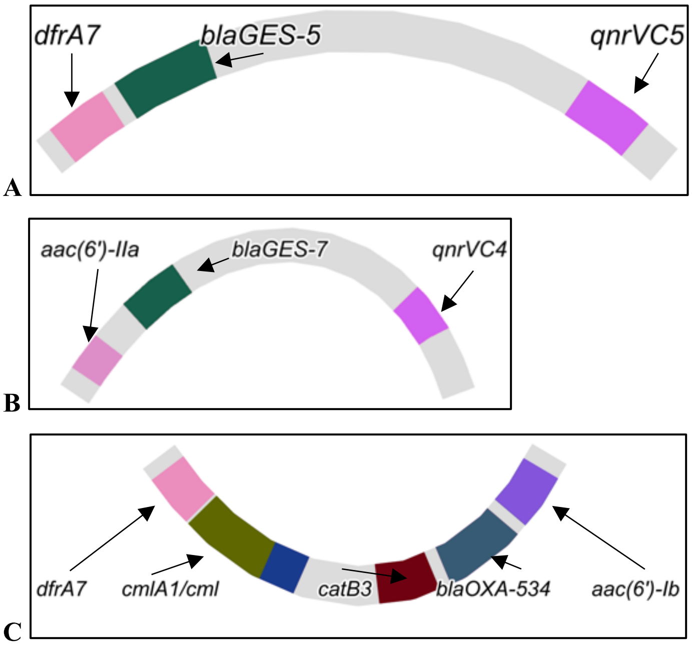


**Figure S5.** Examples of assembly graphs showing multidrug-resistance fragments putatively encoded by class 1 integron gene cassettes in samples A) BH02, B) BH48, and C) BFH19. Contigs were assembled with MEGAHIT [3] (v1.2.8) with parameters --min-contig-len 1000 -m 32000000000. To visualize the co-localization of multiple ARGs with Bandage tool [4], the assembled contigs were converted into assembly graphs using the MEGAHIT command 'megahit_toolkit contig2fastg'. Contigs (putative class 1 integron gene cassettes) carrying multiple ARGs were browsed visually with Bandage tool [4] and ResFinder database [5].

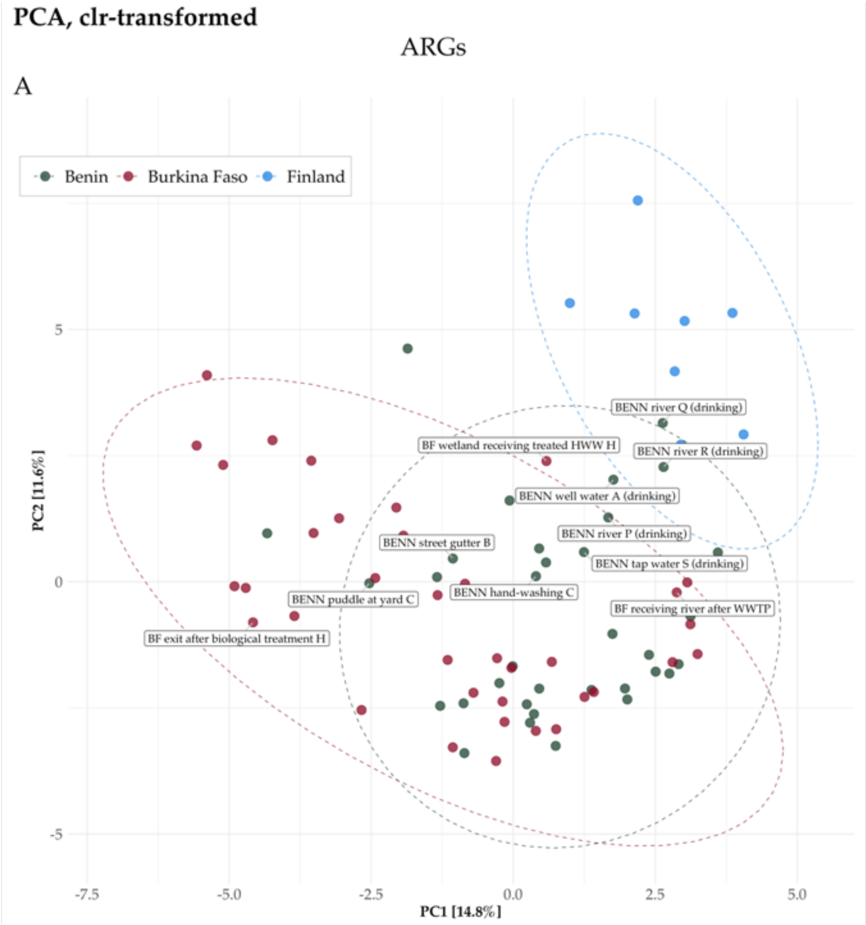
PCA, clr-transformed
ARGs
A
Benin
Burkina Faso
Finland
BENN river Q (drinking)
BF wetland receiving treated HWW H
BENN river R (drinking)
BENN well water A (drinking)
BENN river P (drinking)
BENN street gutter B
BENN tap water S (drinking)
BENN hand-washing C
BENN puddle at yard C
BF receiving river after WWTP
BF exit after biological treatment H
PC2 [11.6%]
PC1 [14.8%]
5
0
-5
-7.5
-5.0
-2.5
0.0
2.5
5.0

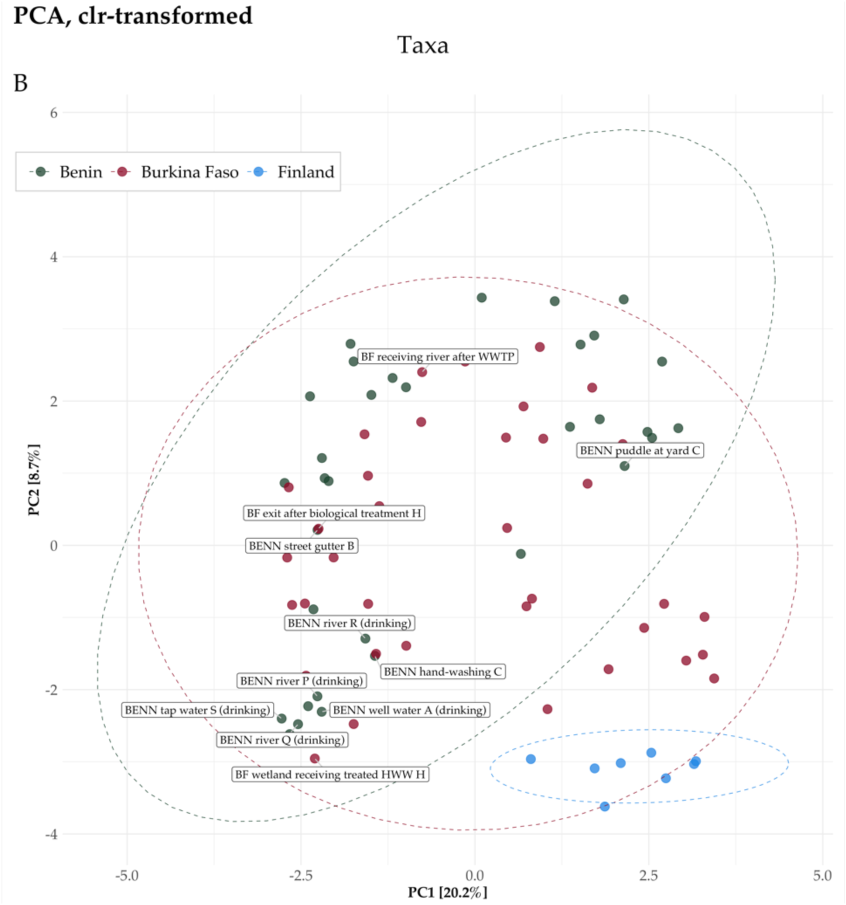
PCA, clr-transformed
Taxa
B
Benin
Burkina Faso
Finland
BF receiving river after WWTP
BENN puddle at yard C
BF exit after biological treatment H
BENN street gutter B
BENN river R (drinking)
BENN hand-washing C
BENN river P (drinking)
BENN tap water S (drinking)
BENN well water A (drinking)
BENN river Q (drinking)
BF wetland receiving treated HWW H
PC2 [8.7%]
PC1 [20.2%]
6
4
2
0
-2
-4
-5.0
-2.5
0.0
2.5
5.0

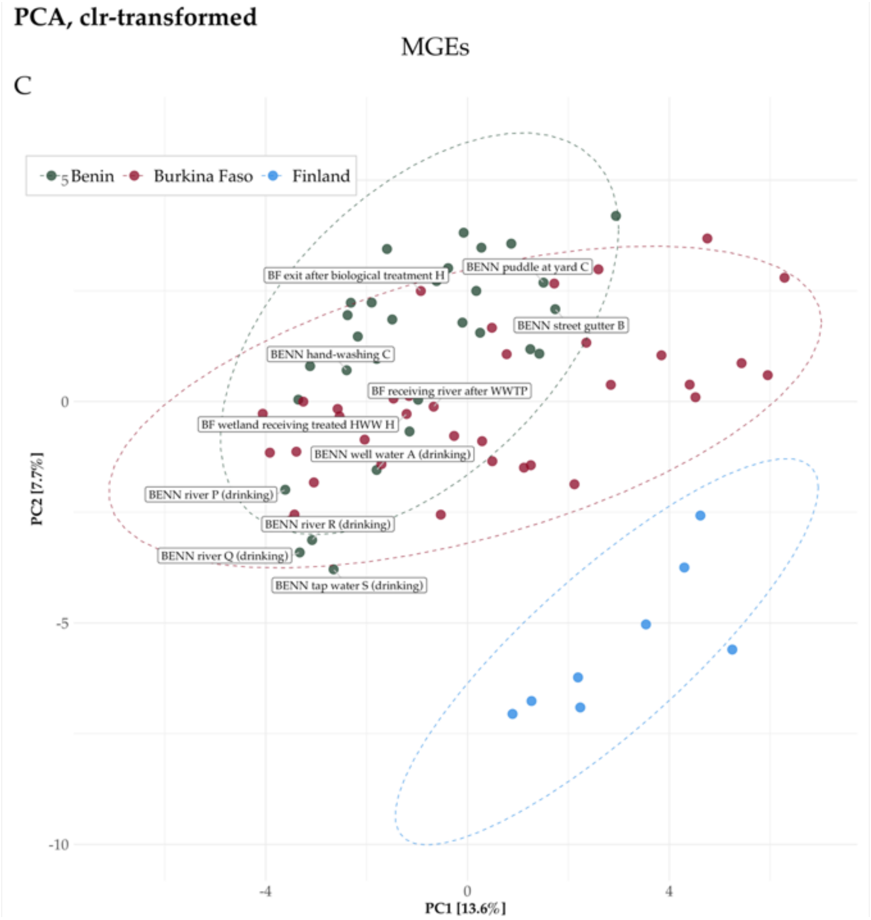


**Figure S6.** Principal components analysis (PCA) showing the significant dissimilarities of A) resistomes, B) taxonomical composition Metaphlan3, and C) mobilomes among all collected (HWW and other water samples) samples from Benin, Burkina Faso, and Finland. Non-HWW, samples are labeled with a text box, while HWW samples are unlabeled. Count data were clr-transformed and visualized with 'microViz' [6] (v0.9.1). Confidence ellipses represent 95 % confidence levels.

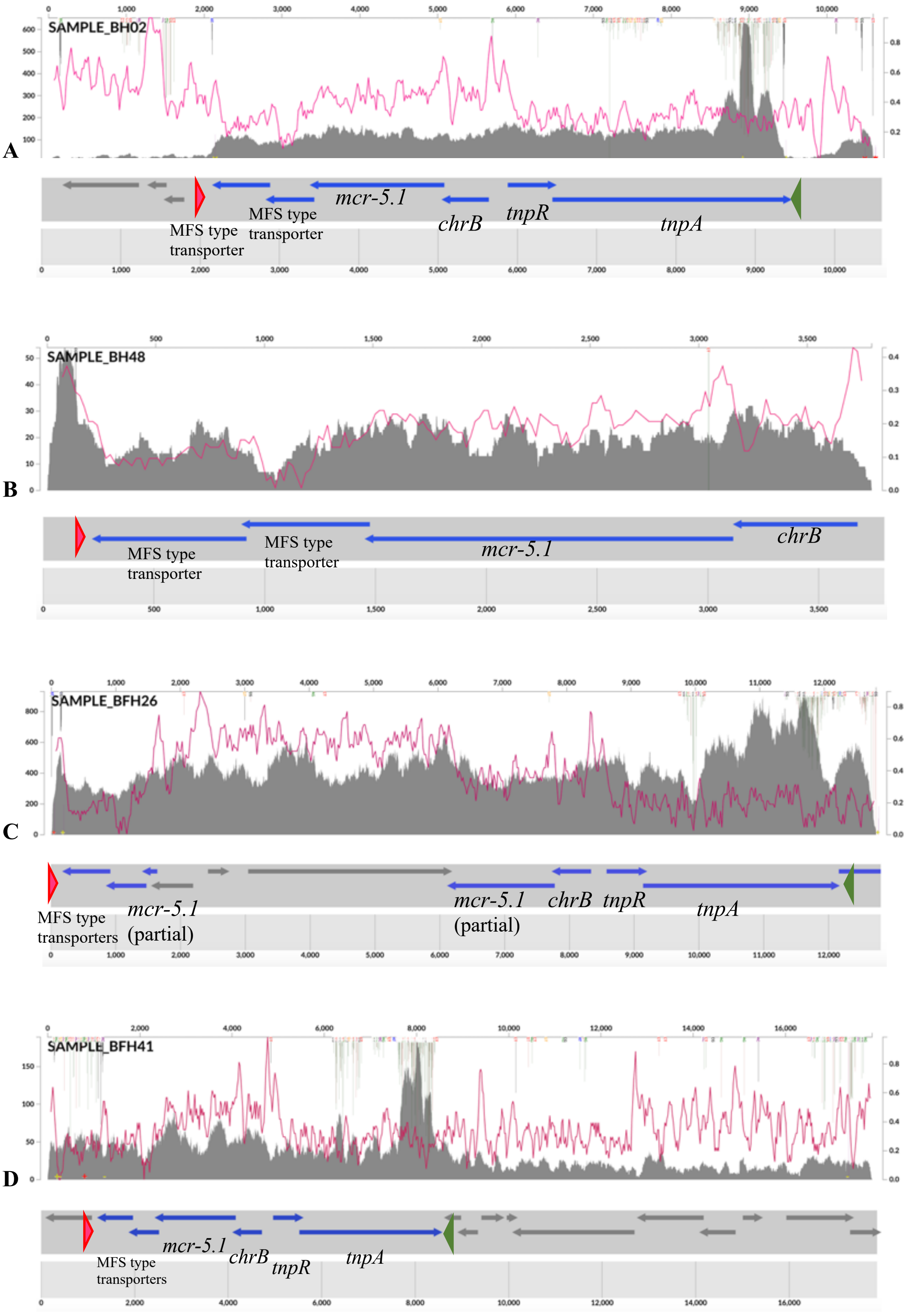
A
SAMPLE_BH02
MFS type transporter
MFS type transporter
mcr-5.1
chrB
tnpR
tnpA
B
SAMPLE_BH48
MFS type transporter
MFS type transporter
mcr-5.1
chrB
C
SAMPLE_BFH26
MFS type transporters
mcr-5.1 (partial)
mcr-5.1 (partial)
chrB
tnpR
tnpA
D
SAMPLE_BFH41
MFS type transporters
mcr-5.1
chrB
tnpR
tnpA

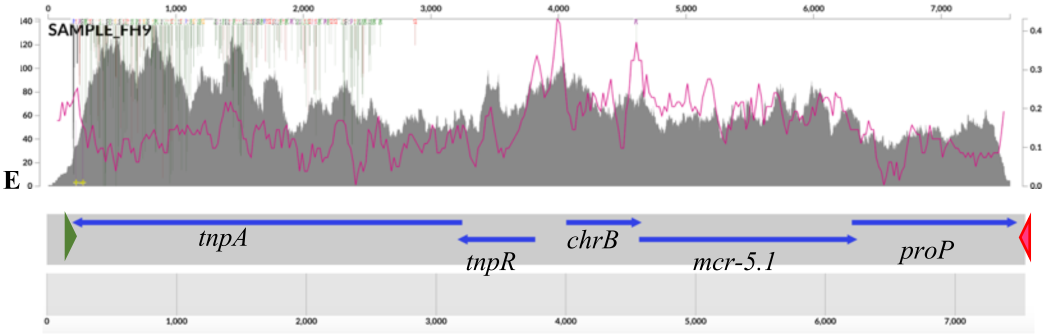


**Figure S7.** The genetic environments of *mcr-5* genes in four different metagenomic samples A) BH02 B) BH48, C) BFH26 D) BFH41, and E) FH9. Metagenomic contigs were assembled with MEGAHIT (v1.2.8) with parameters --min-contig-len 1000 -m 32000000000 [3]. Visualizations of the contigs were created using Anvi'o (v7) metagenomics workflow for single profiles [7]. Blue arrows indicate putative encoding genes. The gene names were determined based on BLAST search results. The recruited reads are shown as a dark grey histogram with an overlay line (pink) denoting the GC content. Single nucleotide variation is shown on top of the panel with colorful vertical lines. Inverse repeat sequences of Tn3-like element sequences (38 bp) are shown with dark pink (IRL: 5' GGGGTCGTCTCAGAAAACGGAAAAAATCGTACGCTAAG 3'[8]) and green triangles (IRR: 5' CTTAGCGTACGATTTTTTCCGTTTTCTAGACGACCCC 3' [8]).

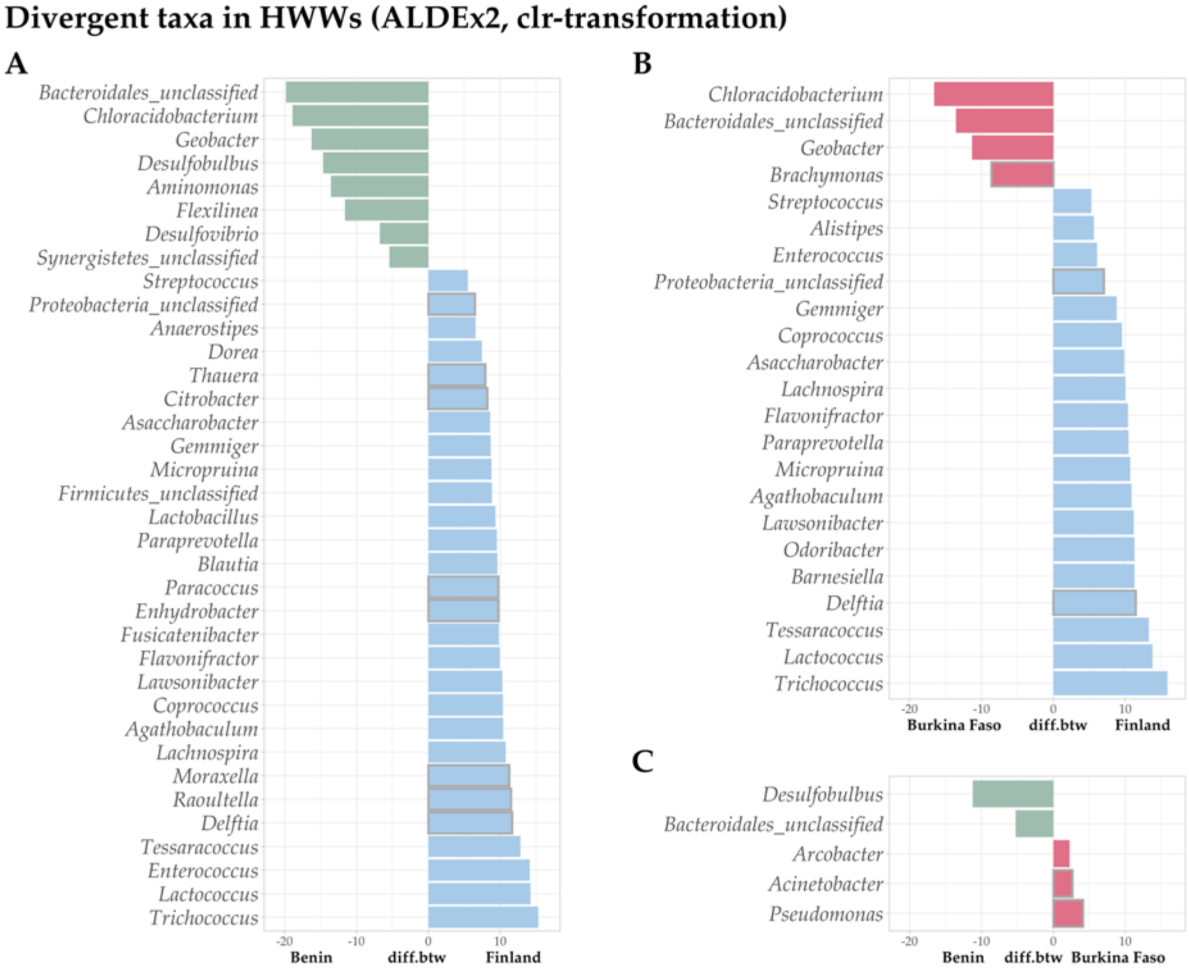

**Figure S8.** Divergent barplot shows the differentially abundant taxa in HWW from Benin, Burkina Faso, and Finland defined by the ALDEx2 tool [9] (v1.28.0). Metaphlan3 count data (obtained by running Metaphlan3 with the parameter '-t rel_ab_w_read_stats') was clr-transformed in the analysis. The pairwise comparisons were A) Benin – Finland, B) Burkina Faso – Finland, and C) Benin – Burkina Faso. Significantly more abundant taxa in one country have positive 'diff.btw' values, while taxa significantly more abundant in the other country have negative 'diff.btw' values. Significantly differentially abundant taxa (BH corrected *p*-value < 0.05) were filtered for visualization to include only taxa with 'diff.btw' value < -5 or > 5, except for the Benin-Burkina Faso, in which all significant results are shown in the plot. All unfiltered results are listed in Tables S8, S9, and S10. Countries are denoted with distinct colors (Benin: green; Burkina Faso: red; Finland: blue). Genera belonging to the phylum *Pseudomonadota* are highlighted with a grey borderline to show the genera considered typical carriers of MGEs such as plasmids and integrons and contribute to the carriage of ARGs [10].

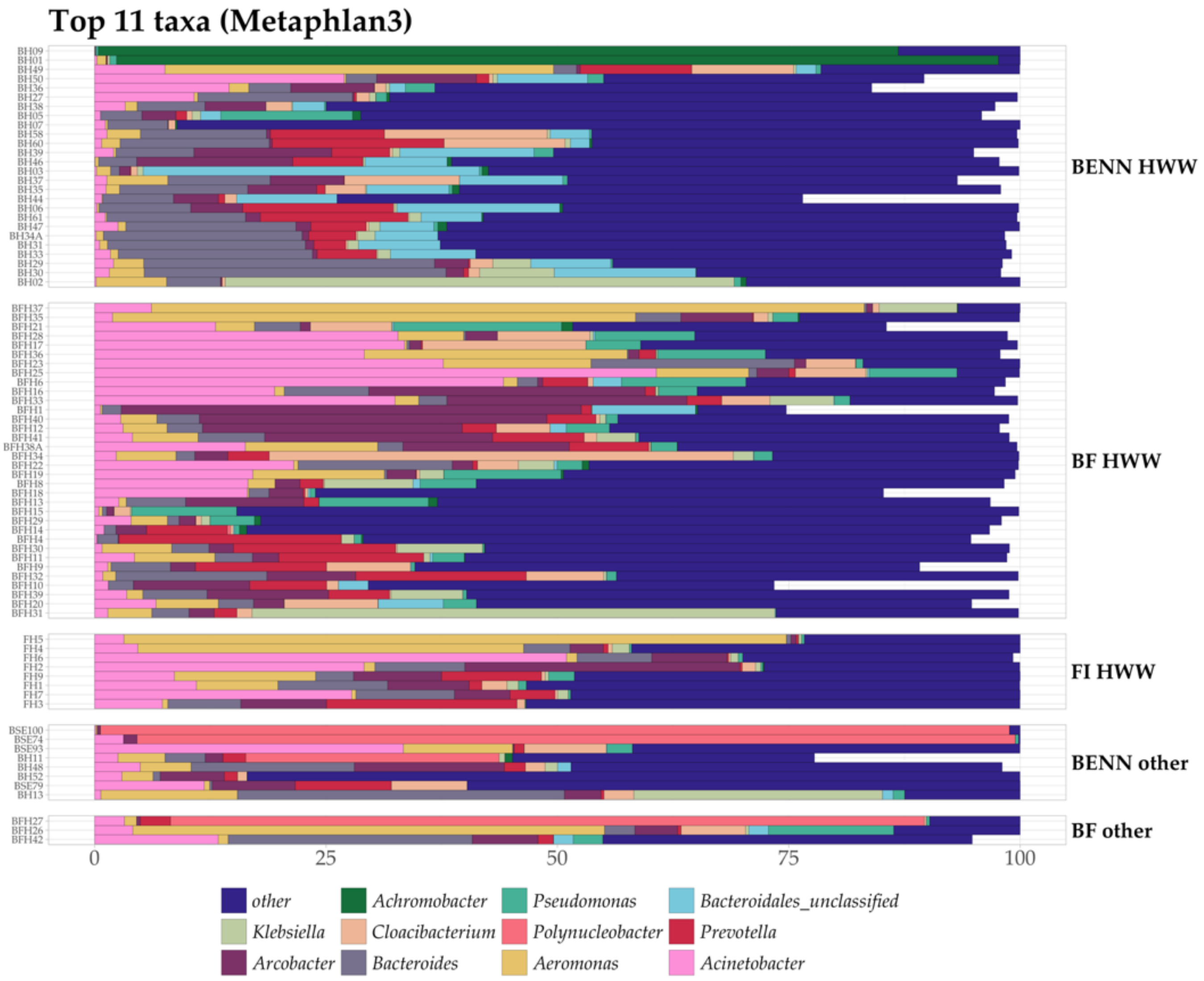


**Figure S9.** Top 11 most abundant genera across all collected samples (HWW and other water samples). Sample codes are on the left, and the sample groups are on the right side of the plot (BENN HWW = HWW from Benin, BF HWW = HWW from Burkina Faso, FI HWW = HWW from Finland, BENN other (from top to bottom) = BENN river P (drinking), BENN river Q (drinking), BENN tap water S (drinking), BENN well water A (drinking),

BENN puddle at yard C, BENN hand-washing C, BENN river R (drinking), BENN street gutter, BF other (from top to bottom) = BF wetland receiving treated HWW H, BF exit after biological treatment H, BF receiving river after WWTP B). Relative abundances were obtained using Metaphlan3 [11] and visualized with 'comp_barplot' from package 'microViz' [6] without any transformation ('tax_transform_for_plot = "identity"'). Samples are ordered according to Bray-Curtis distance.

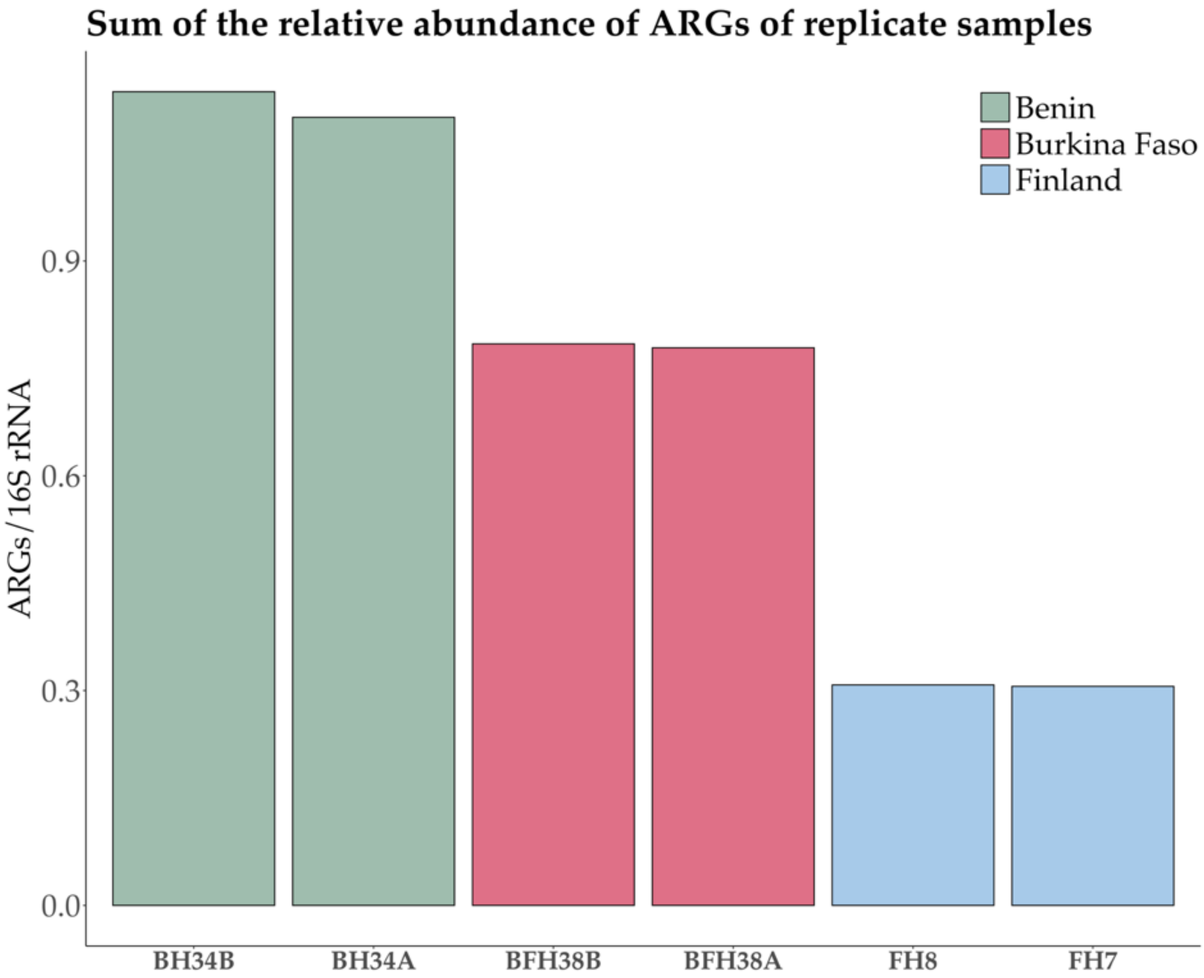


**Figure S10.** The sum of the relative abundance of ARGs in the replicated samples. Sample pairs from Benin (samples BH34 and BH34B), Burkina Faso (samples BFH38A and BFH38B), and Finland (samples FH7 and FH8) were collected from the same septic tanks (Benin and Burkina Faso) or drains (Finland) at the same time.

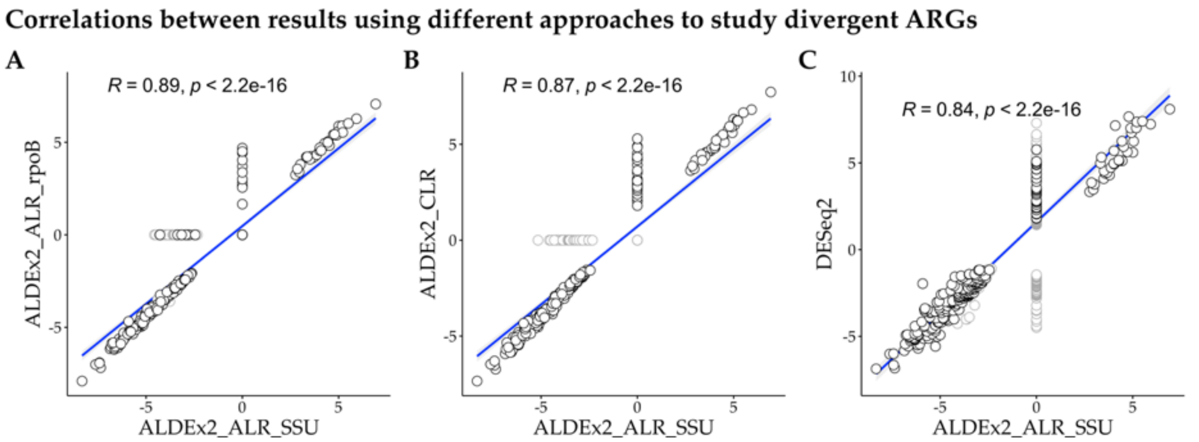


**Figure S11.** Significantly differentially abundant ARGs were studied using different approaches, and the results were compared with each other. The results from ALDEx2 [9] (v1.28.0) using 16S rRNA as the reference were compared to A) those obtained using the *rpoB* gene as a reference, B) those without a reference, and to C) the results from Differential gene expression analysis based on the negative binomial distribution (DESeq2) [12] (v1.36.0). The *rpoB* counts were obtained using the translated sequence reads described above as input for hmmsearch [13] against a *rpoB*-specific HMM profile (pf04563) from the Pfam-A database [14]. Hits or forward reads (R1) were counted. In DESeq2 analysis, ARGs filtered using a variance threshold of 50 were screened for significantly different ARGs between the compare groups after Benjamini-Hochberg adjustment ($p < 0.05$). The DESeq2 algorithm is designed for unnormalized count data from gene expression studies. Thus, to take into account the amount of bacterial DNA in the samples (counts of detected 16S rRNA), the relative abundances were multiplied by $10^5$, and a pseudo count of one was added to avoid zero values unsuitable for integer transformation. HWW from Benin and Finland were included in these comparisons. Insignificant results from one of the two compared approaches were set to zero. Overall, the results conducted using each approach generated similar results considering the specific ARGs but the number of significantly differentially abundant ARGs reported by one method differed from another (zero data points in the correlations). The most drastic difference in the results was seen between ALDEx2 with SSU as reference and DESeq2 (C).

### 3. References


1. Oksanen J, Blanchet G, Friendly M, et al. vegan: Community Ecology Package. https://cran.r-project.org/package=vegan.
2. Kassambara A. ggcorrplot: Visualization of a Correlation Matrix using “ggplot2”. https://cran.r-project.org/package=ggcorrplot. https://cran.r-project.org/package=ggcorrplot
3. Li D, Liu CM, Luo R, Sadakane K, Lam TW. MEGAHIT: an ultra-fast single-node solution for large and complex metagenomics assembly via succinct de Bruijn graph. *Bioinformatics*. 2015;31(10):1674–1676. doi:10.1093/bioinformatics/btv033
4. Wick RR, Schultz MB, Zobel J, Holt KE. Bandage: interactive visualization of de novo genome assemblies. *Bioinformatics*. 2015;31(20):3350–3352. doi:10.1093/bioinformatics/btv383
5. Bortolaia V, Kaas RS, Ruppe E, et al. ResFinder 4.0 for predictions of phenotypes from genotypes. *Journal of Antimicrobial Chemotherapy*. 2020;75(12):3491-3500. doi:10.1093/jac/dkaa345
6. Barnett D, Arts I, Penders J. microViz: an R package for microbiome data visualization and statistics. *J Open Source Softw*. 2021;6(63):3201. doi:10.21105/joss.03201
7. Murat Eren A, Kiefl E, Shaiber A, et al. Community-led, integrated, reproducible multi-omics with anvi’o. *Nat Microbiol*. 2021;6:3-6. doi:10.1038/s41564-020-00834-3
8. Borowiak M, Fischer J, Hammerl JA, Hendriksen RS, Szabo I, Malorny B. Identification of a novel transposon-associated phosphoethanolamine transferase gene, *mcr-5*, conferring colistin resistance in d-

tartrate fermenting *Salmonella enterica* subsp. *enterica* serovar Paratyphi B. *Journal of Antimicrobial Chemotherapy*. 2017;72(12):3317-3324. doi:10.1093/jac/dkx327

9. Fernandes AD, Reid JNS, Macklaim JM, McMurrough TA, Edgell DR, Gloor GB. Unifying the analysis of high-throughput sequencing datasets: Characterizing RNA-seq, 16S rRNA gene sequencing and selective growth experiments by compositional data analysis. *Microbiome*. 2014;2(1). doi:10.1186/2049-2618-2-15

10. Rice EW, Wang P, Smith AL, Stadler LB. Determining Hosts of Antibiotic Resistance Genes: A Review of Methodological Advances. *Environ Sci Technol Lett*. 2020;7(5):282-291. doi:10.1021/acs.estlett.0c00202

11. Beghini F, McIver LJ, Blanco-Míguez A, et al. Integrating taxonomic, functional, and strain-level profiling of diverse microbial communities with biobakery 3. *Elife*. 2021;10. doi:10.7554/eLife.65088

12. Love MI, Huber W, Anders S. Moderated estimation of fold change and dispersion for RNA-seq data with DESeq2. *Genome Biol*. 2014;15:550. doi:10.1186/s13059-014-0550-8

13. Eddy SR. Accelerated Profile HMM Searches. *PLoS Comput Biol*. 2011;7(10):1002195. doi:10.1371/journal.pcbi.1002195

14. El-Gebali S, Mistry J, Bateman A, et al. The Pfam protein families database in 2019. *Nucleic Acids Res*. 2018;47:427-432. doi:10.1093/nar/gky995
